# Development and validation of a novel clinical risk score to predict hypoxemia in children with pneumonia using the WHO PREPARE dataset

**DOI:** 10.1101/2024.08.19.24312238

**Authors:** Rainer Tan, Arjun Chandna, Tim Colbourn, Shubhada Hooli, Carina King, Norman Lufesi, Eric D McCollum, Charles Mwansambo, Joseph L. Mathew, Clare Cutland, Shabir Ahmed Madhi, Marta Nunes, Sudha Basnet, Tor A. Strand, Kerry-Ann O’Grady, Brad Gessner, Emmanuel Addo-Yobo, Noel Chisaka, Patricia L. Hibberd, Prakash Jeena, Juan M. Lozano, William B. MacLeod, Archana Patel, Donald M. Thea, Ngoc Tuong Vy Nguyen, Marilla Lucero, Syed Mohammad Akram uz Zaman, Shinjini Bhatnagar, Nitya Wadhwa, Rakesh Lodha, Satinder Aneja, Mathuram Santosham, Shally Awasthi, Ashish Bavdekar, Monidarin Chou, Pagbajabyn Nymadawa, Jean-William Pape, Glaucia Paranhos-Baccala, Valentina S. Picot, Mala Rakoto-Andrianarivelo, Vanessa Rouzier, Graciela Russomando, Mariam Sylla, Philippe Vanhems, Jianwei Wang, Romina Libster, Alexey W. Clara, Fenella Beynon, Gillian Levine, Chris A Rees, Mark I Neuman, Shamim A. Qazi, Yasir Bin Nisar, World Health Organization PREPARE study group

## Abstract

**Background:** Hypoxemia predicts mortality at all levels of care, and appropriate management can reduce preventable deaths. However, pulse oximetry and oxygen therapy remain inaccessible in many primary care health facilities. We aimed to develop and validate a simple risk score comprising commonly evaluated clinical features to predict hypoxemia in 2-59-month-old children with pneumonia.

**Methods:** Data from 7 studies conducted in 5 countries from the Pneumonia Research Partnership to Assess WHO Recommendations (PREPARE) dataset were included. Readily available clinical features and demographic variables were used to develop a multivariable logistic regression model to predict hypoxemia (SpO_2_<90%) at presentation to care. The adjusted log coefficients were transformed to derive the PREPARE hypoxemia risk score and its diagnostic value was assessed in a held-out, temporal validation dataset.

**Results:** We included 14,509 children in the analysis; 9.8% (n=2,515) were hypoxemic at presentation. The multivariable regression model to predict hypoxemia included age, sex, respiratory distress (nasal flaring, grunting and/or head nodding), lower chest indrawing, respiratory rate, body temperature and weight-for-age z-score. The model showed fair discrimination (area under the curve 0.70, 95% CI 0.67 to 0.73) and calibration in the validation dataset. The simplified PREPARE hypoxemia risk score includes 5 variables: age, respiratory distress, lower chest indrawing, respiratory rate and weight-for-age z-score.

**Conclusion:** The PREPARE hypoxemia risk score, comprising five easily available characteristics, can be used to identify hypoxemia in children with pneumonia with a fair degree of certainty for use in health facilities without pulse oximetry. Its implementation would require careful consideration to limit inappropriate referrals on patients and the health system. Further external validation in community settings in low-and middle-income countries is required.

**KEY MESSAGES:** *What is already known on this topic:*

- Pulse oximetry is unavailable or underutilized in many resource-limited settings in low- and middle-income countries.
- Hypoxemia is a good predictor of mortality and its early identification and further management can reduce mortality.

*What this study adds:*

- The PREPARE hypoxemia risk score was developed using one of the largest and most geographically diverse datasets on childhood pneumonia to date.
- Using age, lower chest indrawing, respiratory rate, respiratory distress and weight-for-age z-score to calculate the PREPARE hypoxemia risk score could help identify children with hypoxemia in settings without pulse oximeters.

*How this study might affect research, practice or policy:*

- This study contributes to the important discussion on how best to identify hypoxemic children in the absence of pulse oximetry.
- Further research is warranted to validate the findings in community settings
- Operationalizing and integrating the score within existing clinical management pathways must be tailored to the setting of implementation.

## INTRODUCTION

Hypoxemia – low blood oxygen levels, defined as an oxygen saturation (SpO_2_) of <90%, is common in children with World Health Organization (WHO) classified pneumonia in low- and middle-income countries (LMICs).^1^ It is a good predictor of preventable deaths,^2-4^ and early identification of hypoxemia and appropriate management can reduce mortality.^5-8^ The availability of pulse oximetry to measure SpO_2_ impacts clinicians’ assessment of illness severity, diagnosis, treatment and need for referral.^6 9^ As such, there is increasing rationale to support universal access to pulse oximetry in primary care settings to address the high burden of pneumonia-related mortality.^10-13^

Although the availability of pulse oximetry is increasing, in part due to increased focus during the COVID-19 pandemic,^10^ access is limited and inequitable and pulse oximetry remains underutilized in many LMIC settings, particularly in the community and primary care health facilities.^14-17^ Barriers to the implementation of pulse oximetry in LMICs include high cost of quality pediatric devices with appropriately sized probes, maintenance, requisite training, battery charging and patient-related factors (e.g. crying and movement), which can be particularly challenging among young children.^18-20^ Furthermore, performing pulse oximetry can take time, especially in agitated children; an important consideration for implementation in health facilities with high case-loads or insufficient staff-to-patient ratios.

In primary care level health facilities without pulse oximetry, where the WHO’s Integrated Management of Childhood Illnesses (IMCI) chartbook^21^ is used to guide the management of sick children, many hypoxemic children are not identified.^6 19^ In health facilities where a pulse oximeter is not available, identifying children at the highest risk of being hypoxemic using clinical signs alone could improve the sensitivity of the IMCI chartbook for identification of children in need of oxygen therapy, hospitalization, or referral. Previous clinical scores to predict hypoxemia were limited in sample size, geographical representation, or included patients already identified as requiring referral based on other clinical features.^22-26^

We aimed to develop and validate a clinical risk score to predict hypoxemia (SpO_2_ <90%) at presentation to a health facility in children aged 2-59 months with pneumonia according to the 2014 WHO IMCI definition (i.e. cough and/or difficulty breathing with fast breathing and/or lower chest indrawing) using the Pneumonia REsearch Partnership to Assess WHO REcommendations (PREPARE) dataset.^21 27^ The goal is not to replace the pulse oximeter, but to guide referral to health facilities where children with suspected hypoxemia can be appropriately managed.

## METHODS

### Study design

We used the WHO PREPARE dataset, to derive and validate a clinical risk score to identify hypoxemia in children 2-59 months with pneumonia. The PREPARE dataset included primary, patient-level data for children 0-59 months with pneumonia, from 45 separate studies conducted from 1994 to 2018, in over 20 countries in Asia, Africa, Latin America, Oceania and North America.^27^ Detailed description of this dataset is found elsewhere.^27^ We adhered to the Transparent Reporting of a Multivariable Prediction Model for Individual Prognosis or Diagnosis (TRIPOD) guidelines.^28^

### Patient and public involvement

The development of the research question was informed by the high rates of pneumonia-related mortality, specifically in children with hypoxemia. Patients were neither advisers in this study nor were they involved in the design, recruitment or conduct of the study. Results of this study will be made publicly available through open-access publication.

### Study population

For the present analysis, included studies that reported SpO_2_ measured at the same time as assessment of clinical symptoms and signs (within 2 hours) and for which pulse oximetry was performed in >=80% of cases. Studies in which fewer than 75% of patients had data for each candidate predictor were excluded. Patients aged 2-59 months with IMCI defined pneumonia (irrespective of SpO_2_ reading) presenting to health facilities were included. Patients with severe pneumonia (defined as cough and/or difficulty breathing with a general danger sign including stridor in a calm child), or complicated severe acute malnutrition were excluded, since they would already have met IMCI criteria for referral.^21 29^

### Outcome

The primary outcome was hypoxemia, defined as a SpO_2_ of <90%,^21, 27^ assessed at the same time as the clinical predictors (symptoms and signs). A sensitivity analysis assessed the development of a model for the prediction of blood oxygen saturation at a SpO_2_ value of <92%. This threshold was selected acknowledging the high risk of mortality in children at this threshold,^13^ and the use of this threshold by various clinical guidelines.^30 31^

### Candidate Predictors

Candidate predictors were selected a priori based on expert knowledge and previous diagnostic value for predicting hypoxemia, identified in the literature,^26^ and considering reliability, feasibility and resources available at typical primary care health facilities in LMICs. The seven candidate predictors were: sex, age, chest indrawing, respiratory rate, axillary temperature, weight-for-age z-score and respiratory distress (composite predictor defined as the presence of either nasal flaring, grunting and/or head nodding). A sensitivity analysis assessed the performance of the model without respiratory distress, with and without wheezing. Wheezing was included in a sensitivity analysis because it is difficult to identify without the use of a stethoscope, often absent or infrequently used in primary care health facilities in LMICs, however included within IMCI.^21 32^ In the analyses without respiratory distress as a candidate predictor, we excluded all patients with signs of respiratory distress. Given the variety of aims and methods among studies in the PREPARE dataset, some clinical predictors may have been measured with knowledge of the outcome status (peripheral oxygen saturation). Duration of cough or difficulty breathing, history of fever, duration of fever, mid-upper arm circumference, weight-for-height z-score, tracheal tugging, heart rate, cyanosis and capillary refill time, were considered as candidate predictors based on potential clinical relevance, but were not included due to high levels of missingness in the data.

### Sample size

We used 75% of the dataset for the development of the score and 25% for validation. Each study dataset was split by reserving the last 25% of enrolled patients for the validation dataset. Such a temporal validation strategy is considered intermediate between internal and external validation.^28^ Using the approach outlined by Riley et al.,^33^ 659 cases of hypoxemia would be required in order to have a sufficient sample size to include the 12 pre-specified parameters (from the 7 candidate predictors) to develop the score (54.9 events per parameter), considering a conservative R^2^ Nagelkerke of 0.15, shrinkage factor of 0.9 and outcome prevalence of 9.8% in the development dataset. The development dataset of 10,884 children contained 1,146 cases of hypoxemia (i.e. 95.5 outcome events per parameter). The temporal validation dataset included 275 outcome events, well above the recommended 100 events often recommended for a robust validation.^34 35^

### Analysis

Pulse oximetry measurements were not adjusted for altitude because no sites were greater than 2,500 meters above sea level.^36^ Continuous predictors were categorized a priori based on currently accepted thresholds in order to maximize interpretability, encourage uptake amongst healthcare providers and policy makers and align with current clinical practices: age: 2-5 months, 6-11 months, 12-59 months; temperature: <35.5, 35.5-37.4, >=37.5°C; respiratory rate: 0-9, 10-19, >=20 breaths/min above IMCI age-specific cut-offs (age 2-11 months >=50 breaths per minute; age 12-59 months >=40 breaths per minute); and weight-for-age z-score: >= -2, < -2 to -3, < -3. Patients with missing data were excluded as multiple imputation was not feasible as it was determined that data were unlikely to be missing at random.^37^

The model was developed using multivariable logistic regression with the least absolute shrinkage and selection operator (LASSO) with L1 regularization. The adjusted model included all parameters and was further adjusted for study. Associations with 95% confidence intervals (CI) for adjusted odds ratios (aOR) that did not cross 1 were considered significant. Discrimination was evaluated by calculating the area under the receiver operating characteristic (AUROC) curve of the model in both the development and validation datasets. Calibration was evaluated using calibration plots (observed risk on y-axis, predicted risk on the x-axis) in the validation dataset.

In order to convert the model into a simple clinical risk score, the regression coefficient of each retained parameter with a statistically significant adjusted odds ratio (CI did not overlap with 1.0) was rounded to the nearest 0.5 and then doubled.^38 39^ Sensitivity, specificity, likelihood ratios, miss rate (proportion of patients with hypoxemia not identified), false discovery rate (proportion of patients inappropriately identified as having hypoxemia among all patients identified as having hypoxemia) and proportion of children identified as being hypoxemic at each of the score’s cut offs was evaluated in the temporal validation dataset.

Recognizing that the appropriate threshold for referring a child with suspected hypoxemia would be context dependent, a decision curve analysis compared the clinical utility (net benefit) of the model and score across a range of clinically-plausible referral thresholds. The net benefit quantifies the trade-off between true positives (hypoxemic children predicted to have hypoxemia) and false positives (non-hypoxemic children incorrectly predicted to have hypoxemia), weighted according to the relative cost of a false positive (threshold probability), which would vary in different contexts.

The threshold probability reflects the cut-off (predicted probability of hypoxemia) above which a given intervention (in this case, referral) might be considered.

All analyses were performed using STATA version 16.^40^

## RESULTS

### Baseline characteristics

The PREPARE dataset included 294,968 children from 45 studies, of which 14,509 children from 7 studies met inclusion criteria and were included in this analysis (figure 1). Four studies were randomized controlled trials, and three were prospective cohort studies (table 1). All studies included outpatients presenting to hospitals, and one study also included patients presenting to health centers.^41^ Among the 7 studies included in this analysis, patients were recruited from 5 countries: Australia, Gambia, India, Malawi and Nepal (table 1). Of the 14,509 patients with pneumonia, 9.8% had a SpO_2_ less than 90% at presentation (table 2).

**Table 1:** Characteristics of studies included in the development and validation of the PREPARE hypoxemia risk score.

| Study | Study design | Location | Age | Original inclusion criteria from the source studies | Original exclusion criteria from the source studies | Sample size<sup>a</sup>, n/N (%) | Hypoxemia (SpO<sub>2</sub> <90%) n/N (%) |
| --- | --- | --- | --- | --- | --- | --- | --- |
| Basnet, 2012<sup>42</sup> | Randomized controlled trial | Kathmandu, Nepal | 2-35 months | - Cough for <14 days<br>- Difficulty breathing ≤72 hours with presence of lower chest indrawing on examination | - Children with recurrent wheezing, heart disease, other severe illness, severe malnutrition, dehydration, hemoglobin <70 g/L, chronic cough, effusion on chest x-ray, or history of documented tuberculosis. | 378/641 (59.0%) | 251/378 (76.4%) |
| Cutts, 2005<sup>41</sup> | Randomized controlled trial | Eastern Gambia | 40-364 days | Children with WHO-defined pneumonia of any severity<sup>b</sup> | - Intent to move out of the study area within 4 months<br>- Previous or uncertain receipt of diphtheria-pertussis-tetanus/*Haemophilus influenzae* type b (DPT/Hib) or DPT vaccine<br>- Inclusion in a previous vaccine trial<br>- Serious chronic illness | 433/1,716 (25.2%) | 22/433 (5.1%) |
| Lazzerini, 2016<sup>43</sup> | Prospective cohort | Malawi | 2-59 months | - Children with WHO-defined pneumonia of any severity<sup>b</sup> | None | 7,902/16,475 (48.0%) | 737/7,902 (9.3%) |
| Mathew, 2015<sup>44</sup> | Prospective cohort | Chandigarh, India | 2-59 months | - Children with WHO-defined pneumonia of any severity<sup>b</sup> | - Duration of illness >7 days<br>- Antibiotics for >24 hours<br>- Previous hospitalization within the preceding 30 days<br>- Children with wheeze who received a single dose of bronchodilator and whose symptoms disappeared | 1,132/2,400 (47.2%) | 134/1,132 (11.8%) |
| McCollum, | Prospective cohort | Mchinji and Lilongwe | 2-59 months | - Children with WHO-defined pneumonia of any severity<sup>b</sup> | None | 4,322/6,764 (63.9%) | 274/4,322 (6.3%) |
| 2017<sup>45</sup> |  | Districts, Malawi |  |  |  |  |  |
| O'Grady, 2012<sup>46</sup> | Randomized controlled trial | Central Australia | 2-59 months | - Children with WHO-defined pneumonia of any severity<sup>b</sup> | Children with wheezing and chronic conditions | 26/147 (17.7%) | 0.0% (0/26) |
| Wadhwa, 2013<sup>47</sup> | Randomized controlled trial | New Delhi, India | 2-24 months | - Children with WHO-defined pneumonia of any severity<sup>b</sup><br>- Crepitations on auscultation | - Need for mechanical ventilation or inotropic medications<br>- Any other serious underlying medical condition | 316/550 (57.5%) | 0.9% (3/316) |

<sup>a</sup> Sample size included in the present analysis over the sample size from the original study
<sup>b</sup> WHO-defined pneumonia of any severity: presence of age-specific fast breathing, lower chest indrawing, or danger signs, in children with a cough or difficulty breathing

**Table 2.** Baseline characteristics of children with pneumonia (fast breathing and/or chest indrawing) with pulse oximetry measurements and complete data on all candidate predictors (n=14,509)

| Characteristic | Total (n=14,509) | Development dataset (n=10,884) | Validation dataset (n=3,625) |
| --- | --- | --- | --- |
| **Age<sup>a</sup>** |  |  |  |
| 2-5 months | 4,298 (29.6%) | 3,400 (31.2%) | 898 (24.8%) |
| 6-11 months | 3,663 (25.2%) | 2,830 (26.0%) | 833 (23.0%) |
| 12-59 months | 6,548 (45.1%) | 4,654 (42.8%) | 1,894 (52.2%) |
| **Sex** |  |  |  |
| Male | 8,193 (56.5%) | 6,126 (56.3%) | 2,068 (57.1%) |
| Female | 6,316 (43.5%) | 4,759 (43.7%) | 1,557 (42.9%) |
| **Pneumonia classification** |  |  |  |
| Fast breathing only | 5,519 (38.0%) | 4,052 (37.2%) | 1,467 (40.5%) |
| Lower chest indrawing | 8,990 (62.0%) | 6,832 (62.8%) | 2,158 (59.6%) |
| **Any sign of respiratory distress (nasal flaring, grunting or head nodding)** |  |  |  |
| No | 7,942 (54.7%) | 5,930 (54.5%) | 2,012 (55.5%) |
| Yes | 6,567 (45.3%) | 4,954 (45.5%) | 1,613 (44.5%) |
| **Respiratory rate** |  |  |  |
| < age-adjusted tachypnea threshold<sup>b</sup> | 163 (1.1%) | 124 (1.1%) | 39 (1.1%) |
| 0-9 breaths/min above cut-off | 6,164 (42.5%) | 4,600 (42.3%) | 1,563 (43.1%) |
| 10-19 breaths/min above cut-off | 5,333 (36.8%) | 3,994 (36.7%) | 1,340 (37.0%) |
| >=20 breaths/min above cut-off | 2,849 (19.6%) | 2,166 (19.9%) | 683 (18.8%) |
| **Body temperature** |  |  |  |
| Normal (>=35.5°C to 37.5°C) | 9,402 (64.8%) | 6,908 (63.5%) | 2,494 (68.8%) |
| >=37.5°C | 4,949 (34.1%) | 3,854 (35.4%) | 1,095 (30.2%) |
| <35.5°C | 158 (1.1%) | 122 (1.1%) | 36 (1.0%) |
| **Weight for age z-score** |  |  |  |
| >=-2 | 11,767 (81.1%) | 8,777 (80.6%) | 2,990 (82.5%) |
| <-2 to -3 | 1,777 (12.2%) | 1,364 (12.5%) | 413 (11.4%) |
| <-3 | 965 (6.7%) | 743 (6.8%) | 222 (6.1%) |
| **Oxygen saturation (SpO2)** |  |  |  |
| >=90% | 13,088 (90.2%) | 9,738 (89.5%) | 3,350 (92.4%) |
| <90% | 1,421 (9.8%) | 1,146 (10.5%) | 275 (7.6%) |

<sup>a</sup> Some studies did not include patients in the whole 2-59 month age range; <sup>b</sup> IMCI chartbook threshold for tachypnea: age 2-11 months >=50 breaths per minute; age 12-59 months >=40 breaths per minute

**Figure 1:**
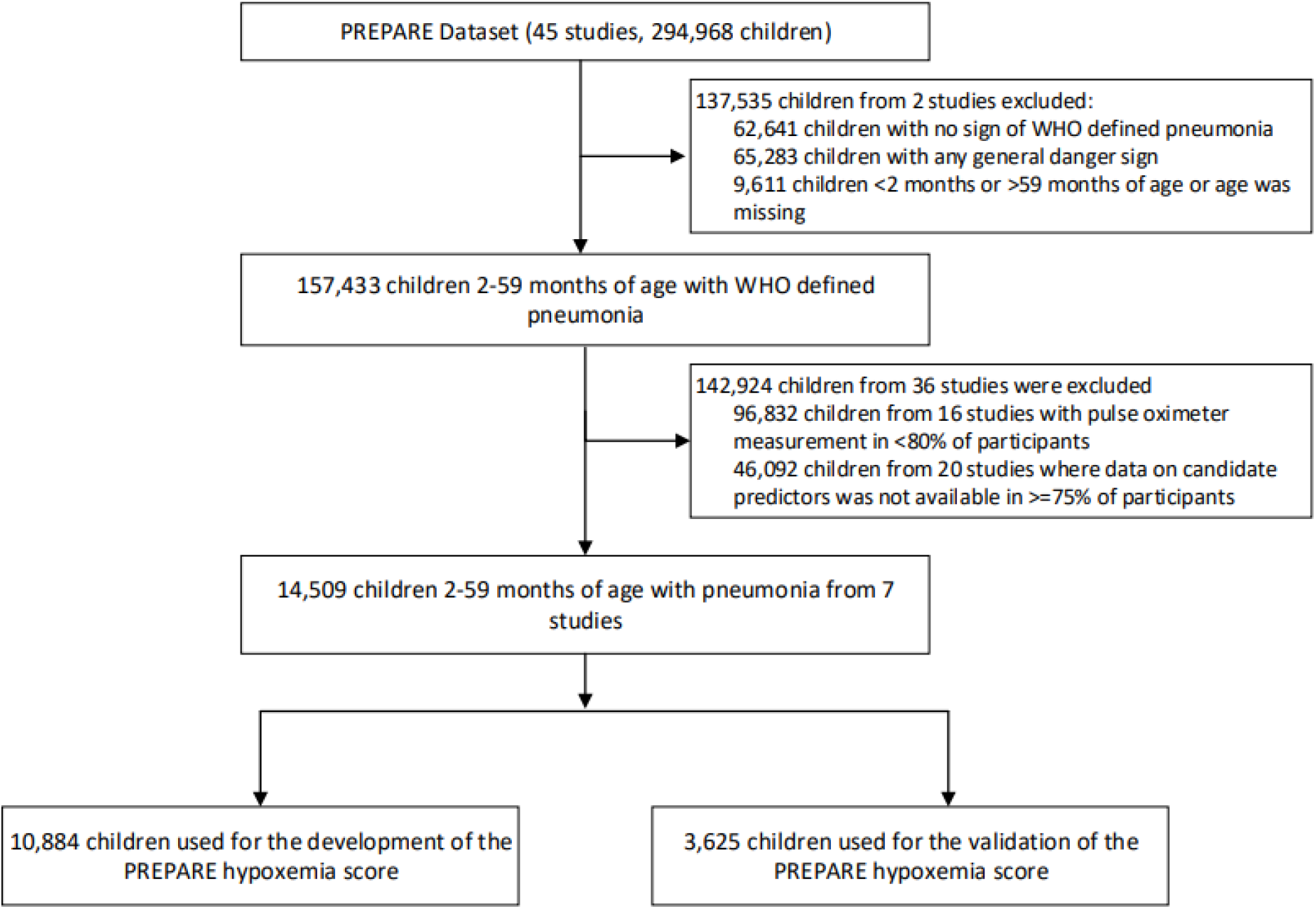
Flow diagram of PREPARE study dataset describing which datasets were excluded/included, the split between development and validation datasets and proportion of patients meeting the primary outcome (SpO2 <90%) Danger signs were defined by the IMCI^8^ chartbook (i.e., inability to drink, lethargy or unconsciousness, convulsions, vomiting everything, or stridor in a calm child).

Baseline characteristics between the development (n=10,884, 75%) and validation dataset (n=3,625, 25%) were similar (table 2). Within the total dataset (n=14,509), 54.9% of children were aged less than 1 year, there were more male patients than female patients, 62.0% of children had lower chest indrawing, 34.1% had a temperature of equal to or more than 37.5°C and 6.7% had a very low weight-for-age z-score (< -3).

### Model development

The predictors most strongly associated with hypoxemia (SpO_2_ <90%) were respiratory rate >/= 20 breaths/minute above the IMCI age-adjusted cut-off (aOR 2.41, 95% CI 1.25 to 4.64), any sign of respiratory distress (aOR 2.21, 95% CI 1.90 to 2.58), weight for age z-score <-3 (aOR 1.96, 95% CI 1.56 to 2.42) and lower chest indrawing (aOR 1.62, 95% CI 1.35 to 1.95) (table 3). None of the 12 candidate parameters were eliminated by the LASSO penalty during model development and hence, all 7 candidate predictors were retained in the final model.

**Table 3.**
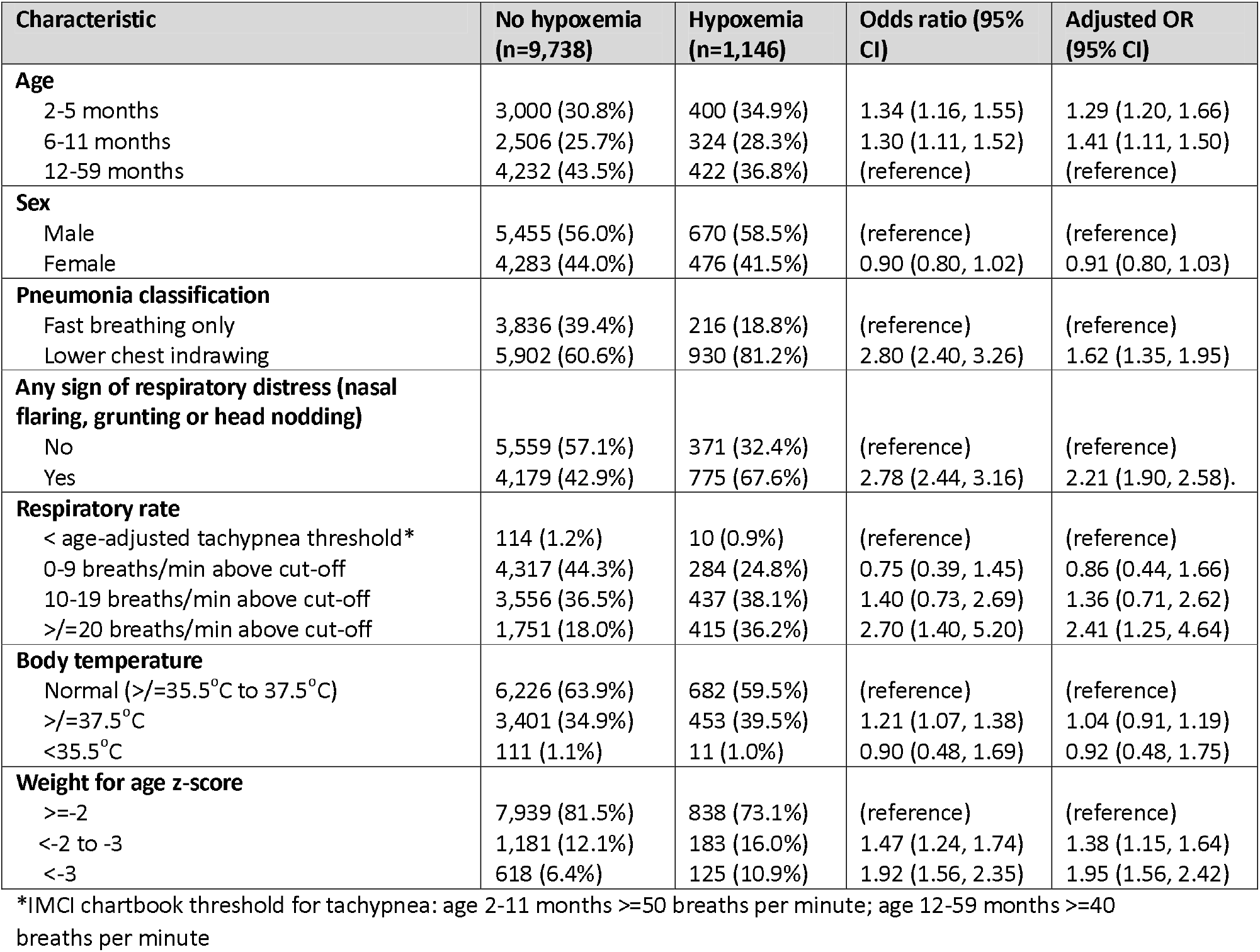
Multivariable regression model for predicting hypoxemia in the development dataset (n=10,884)

### Model performance

The PREPARE hypoxemia clinical prediction model had an AUROC of 0.70 (95% CI 0.69 to 0.72) in the development dataset and an AUROC of 0.70 (95% CI 0.68 to 0.73) in the temporal validation dataset (figure 2). Calibration of the PREPARE hypoxemia prediction model in the validation dataset is illustrated in figure 3.

**Figure 2.**
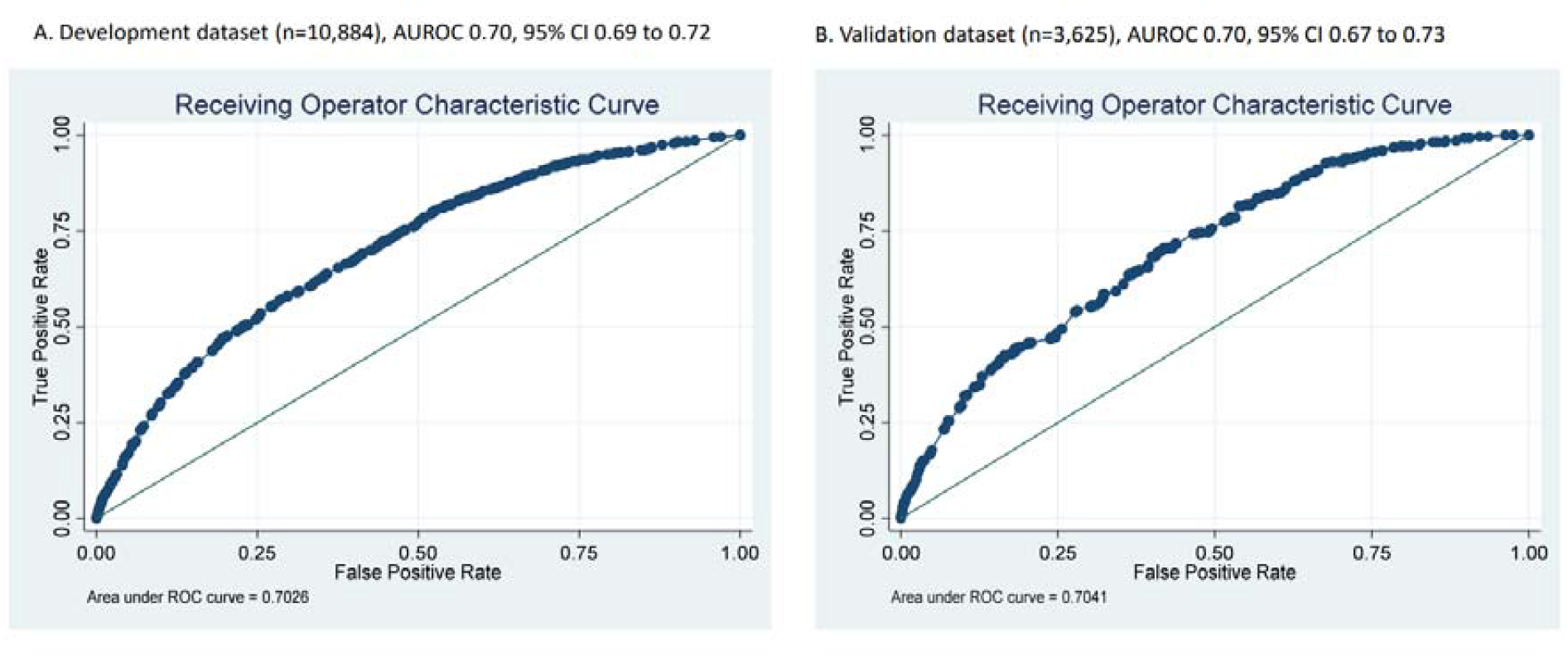
Receiver operating characteristic curve for the PREPARE hypoxemia clinical prediction model for children 2–59 months of age with pneumonia

**Figure 3:**
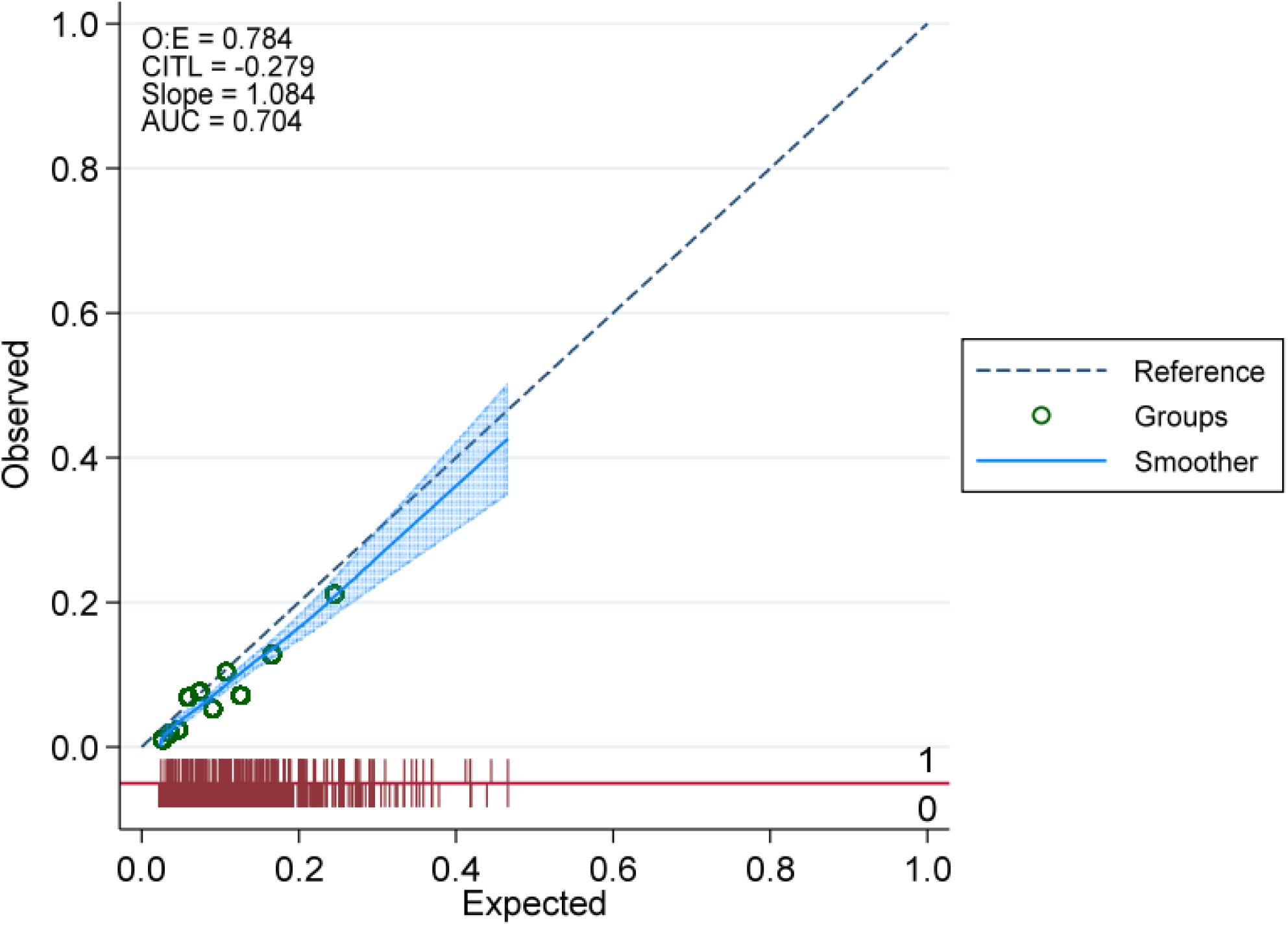
Calibration plot of the PREPARE hypoxemia clinical prediction model in the validation dataset Reference (dashed line) indicates perfect calibration; Lowess (blue line) indicates calibration slope; Red rug plots indicate the distribution of predicted risk of participants with hypoxemia (above the line) and those without hypoxemia (below the line), E:O = expected to observed events ratio; CITL = calibration-in-the-large; AUC = area under the receiver operating characteristic curve.

When converted into the PREPARE hypoxemia risk score (table 4), the retained predictors included age 2-11 months (+1 point), lower chest indrawing (+1 points), respiratory rate >/=20 breaths/min above age-adjusted tachypnea threshold (+2 points), any sign of respiratory distress (+2 points) and weight for age z-score <-2 (+1 point), with the score ranging from 0-7 points (table 4 and 5). Sensitivity, specificity, positive and negative likelihood ratios, miss rate, false discovery rate and the proportion of children identified as hypoxemic are included for each score cut-off (table 5).

**Table 4:** Components of the PREPARE hypoxemia risk score in the development dataset (n=10,884)

| Factor | Adjusted log coefficient | PREPARE hypoxemia risk score<sup>a</sup> |
| --- | --- | --- |
| **Age** |  |  |
| 2-5 months | 0.26 | +1 |
| 6-11 months | 0.34 | +1 |
| 12-59 months | -- | --- |
| **Pneumonia classification** |  |  |
| Fast breathing only | -- | --- |
| Lower chest indrawing | 0.49 | +1 |
| **Any sign of respiratory distress (nasal flaring, grunting or head nodding)** |  |  |
| No | -- | -- |
| Yes | 0.79 | +2 |
| **Respiratory rate** |  |  |
| < age-adjusted tachypnea threshold<sup>b</sup> | -- | --- |
| 0-9 breaths/min above cut-off | -0.15 | --- |
| 10-19 breaths/min above cut-off | 0.31 | --- |
| $\geq 20$ breaths/min above cut-off | 0.88 | +2 |
| **Weight for age z-score** |  |  |
| $\geq -2$ | -- | --- |
| <-2 to -3 | 0.32 | +1 |
| <-3 | 0.66 | +1 |

<sup>a</sup>Regression coefficients of each retained parameter with a statistically significant adjusted odds ratio (confidence interval (CI) did not overlap with 1.0) was rounded to the nearest 0.5 and then doubled to derive the points contributing to the PREPARE hypoxemia risk score.
<sup>b</sup>IMCI chartbook threshold for tachypnea: age 2-11 months $\geq 50$ breaths per minute; age 12-59 months $\geq 40$ breaths per minute

**Table 5.** Performance of the PREPARE hypoxemia risk score at each cut-off in the validation dataset (n=3,625)

| Score | Hypoxemia, n/N (%) | Sensitivity (95% CI)* | Specificity (95% CI)* | Positive Likelihood Ratio (95% CI)* | Negative Likelihood Ratio (95% CI)* | Miss rate† % (n/N) | False discovery rate‡ % | Proportion referred^ % (n/N) |
| --- | --- | --- | --- | --- | --- | --- | --- | --- |

|  |  |  |  |  | CI)* |  | (n/N) |  |
| --- | --- | --- | --- | --- | --- | --- | --- | --- |
| 0 | 6/450<br>(1.3%) | 100.0<br>(98.7,<br>100.0) | 0.0 (0.0,<br>0.1) | 1.00 | --- | 0% (0/275) | 92.4%<br>(3350/3625) | 100%<br>(3625/3625) |
| 1 | 21/781<br>(2.7%) | 97.8<br>(95.3,<br>99.2) | 13.3(12.1,<br>14.5) | 1.13 (1.10,<br>1.15) | 0.16 (0.07,<br>0.36) | 2.2%<br>(6/275) | 91.5%<br>(2906/3175) | 87.6%<br>(3175/3625) |
| 2 | 44/545<br>(8.1%) | 90.2<br>(86.0,<br>93.4) | 35.9 (34.3,<br>37.6) | 1.41 (1.34,<br>1.47) | 0.27 (0.19,<br>0.39) | 9.8%<br>(27/275) | 89.6%<br>(2146/2394) | 66.0%<br>(2394/3625) |
| 3 | 56/737<br>(7.6%) | 74.2<br>(68.6,<br>79.3) | 50.9 (49.2,<br>52.6) | 1.51 (1.40,<br>1.63) | 0.51 (0.41,<br>0.62) | 25.8%<br>(71/275) | 89.0%<br>(1645/1849) | 51.0%<br>(1849/3625) |
| 4 | 67/689<br>(9.7%) | 53.8<br>(47.7,<br>59.8) | 71.2 (69.7,<br>72.8) | 1.87 (1.66,<br>2.11) | 0.65 (0.57,<br>0.74) | 46.2%<br>(127/275) | 86.7%<br>(964/1112) | 30.7%<br>(1112/3625) |
| 5 | 45/284<br>(15.8%) | 29.5<br>(24.1,<br>35.2) | 89.8 (88.7,<br>90.8) | 2.89 (2.34,<br>3.55) | 0.79 (0.73,<br>0.85) | 70.5%<br>(194/275) | 80.9%<br>(342/423) | 11.7%<br>(423/3625) |
| 6 | 28/119<br>(23.5%) | 13.1 (9.3,<br>17.7) | 96.9 (96.3,<br>97.5) | 4.26 (2.96,<br>6.10) | 0.90 (0.86,<br>0.94) | 86.9%<br>(239/275) | 74.1%<br>(103/139) | 3.8%<br>(139/3625) |
| 7 | 8/20<br>(40.0%) | 2.9 (1.3,<br>5.7) | 99.6 (99.4,<br>99.8) | 8.12 (3.35,<br>19.70) | 0.97 (0.95,<br>0.99) | 97.1%<br>(267/275) | 60.0% (12/20) | 0.6%<br>(20/3625) |

\*Calculated at ≥ each respective cut-off
†Miss rate (also known as false negative rate = False Negative/(True Positive + False Negative)
‡False discovery rate = False Positive/(False Positive+True Positive)
^Referral used as a proxy for identification of hypoxemia, assuming that all children identified as at risk of hypoxemia would be referred

In the absence of pulse oximetry, the decision curve analysis suggests that in contexts where it might be acceptable to identify (and refer) up to six non-hypoxemic children for each hypoxemic child (up to a threshold probability of ∼17%) it would be optimal to use a cut-off of >/= 5 to guide referral decision for further management, whereas in contexts in which greater than 1-in-7 true positives are required, a cut-off of >/= six would be preferable (figure S1). The PREPARE hypoxemia model had an equivalent or higher net benefit than the PREPARE hypoxemia risk score (at any cut-off) across all threshold probabilities.

### Prediction of hypoxemia at SpO_2_ threshold of < 92%

In our sensitivity analysis using the same dataset, 15.7% (2,281/14,509) children had SpO2 <92%. The proportion with SpO_2_ <92% was slightly lower in the validation dataset (14.2%, 514/3,625) compared to the development dataset (16.2%, 1,767/10,884). The predictive value of predictors in predicting SpO_2_ <92% compared to SpO_2_ <90% were similar (table S1, table 3).

The PREPARE SpO_2_ <92% clinical prediction model had a similar AUROC (0.67, 95% CI 0.65 to 0.70) compared to the PREPARE hypoxemia (SpO_2_ <90%) prediction model (AUROC 0.70, 95% CI 0.68 to 0.73) (figure S2). The calibration plot for this model is shown in figure S3 and the performance of the risk score at each cut-off is presented in table S3.

The PREPARE SpO_2_ <92% risk score integrates 5 clinical predictors for a total score of 6 points: Age 6-11 months (+1 point), lower chest indrawing (+1 point), respiratory rate >/= 20 breaths/min above age-adjusted tachypnea threshold (+2 points), any sign of respiratory distress (+1 point) and weight-for-age z-score <-3 (+1 point) (table S2).

### PREPARE hypoxemia risk score without respiratory distress

When excluding respiratory distress as a candidate predictor, the dataset was larger with 15,592 children from 14 studies. Five studies were prospective cohorts, 7 were randomized controlled trials, one was a retrospective cohort and one was a prospective case series (table S4). Among the 14 studies included in this analysis, patients were recruited from 23 countries from North, Central and South America, Africa, Asia and Oceania (table S4). Of the 15,592 patients with pneumonia, 16.1% had a SpO_2_ <90% at presentation (table S5). Baseline characteristics were similar in the development and validation datasets (table S5) and did not differ from the primary dataset including respiratory distress. Without respiratory distress, lower chest indrawing (aOR 4.44, 95% CI 3.83 to 5.15) contributed more towards the prediction of hypoxemia (table S6). None of the 11 candidate parameters were eliminated by the LASSO penalty during model development.

The PREPARE hypoxemia clinical prediction model excluding respiratory distress had a higher AUROC than the model that included respiratory distress as a candidate predictor (AUROC 0.78 vs. 0.70) (figure S4). The calibration plot of this model is shown in figure S5.

The final PREPARE hypoxemia risk score when excluding respiratory distress integrated 4 clinical predictors: age 2-11 months (+1 point), lower chest indrawing (+3 points), respiratory rate >=20 breaths/min above age-adjusted tachypnea cut-off (+1 point) and weight for age z-score <-2 (+1 point) for a maximum total score of 6 points (table S7).

### PREPARE hypoxemia risk score with wheezing

Using the same methodology as the primary analysis (i.e., with respiratory distress), including wheezing as a candidate predictor, resulted in a slightly smaller dataset of 13,792 children from 7 studies (table S9). In this dataset, 9.9% had a SpO_2_ of less than 90% at presentation (table S10). Baseline characteristics in these development and validation datasets were similar (table S10) and did not differ from the primary dataset, which excluded wheezing. Wheezing had an adjusted odds ratio of 1.28 (95% CI 1.11 to 1.47), sensitivity of 37.6% and specificity of 73.2% (table S11) for identification of hypoxemia. None of the 12 candidate parameters were eliminated by the LASSO penalty during model development.

The PREPARE hypoxemia clinical prediction model that included wheezing performed similarly to the model without wheezing in the validation datasets (AUROC 0.72 versus 0.70) (figure S6). The calibration plot of the PREPARE hypoxemia clinical prediction model, including wheezing is shown in figure S3. The performance of the PREPARE hypoxemia risk score including wheezing at each cut-off is presented in table S10.

The final PREPARE hypoxemia risk score when including wheezing integrated five clinical parameters: age 2-11 months (+1 points), chest indrawing (+1 point), respiratory distress (+2 point), respiratory rate >=20 breaths/min above cut-off (+2 points) and weight for age z-score <-2 (+1 point) for a maximum total score of 7 points (table S12). Wheezing was not retained in the risk score as the adjusted log coefficient was 0.24. A comparison of all risk score weights from the main analysis and sensitivity analyses can be found in table S13.

## DISCUSSION

We developed the PREPARE hypoxemia risk score from one of the largest and most diverse pneumonia datasets, including over 14,500 childhood pneumonia cases (of which 1,421 had hypoxemia) from 7 studies in 5 countries and in 14 studies in 23 countries within a sensitivity analysis. The components of the score include age, respiratory distress, respiratory rate, lower chest-indrawing and weight-for-age z-score. The risk score demonstrated fair discrimination in predicting hypoxemia (SpO_2_ <90%) in children with pneumonia without danger signs or other indications for referral.

Previous studies predicting hypoxemia included fewer patients from more restricted geographical settings.^8 22-26 48-51^ The use of very large and diverse datasets reduces instability of individual predictions, limits overfitting and provides the opportunity for generalizability to a wide range of settings.^52 53^ A model developed from a dataset with a similar number of patients, though exclusively evaluated in outpatient clinics from Bangladesh and Malawi, had a higher AUROC (0.79) compared to the PREPARE hypoxemia risk score (0.70), but had a similar AUROC to the PREPARE hypoxemia risk score excluding respiratory distress (0.78).^22^ However, this study and many others included patients with IMCI danger signs in whom hospitalization or referral would already be recommended by existing guidelines.^21 22 29^ IMCI danger signs were retained in these prior analyses to evaluate their performance in hypoxemia prediction models alongside other candidate parameters and these studies showed IMCI danger signs added little to no predictive value for hypoxemia.^21^ While excluding such patients in our study limits our ability to comment on the value of IMCI danger signs in hypoxemia prediction models it aligns more closely with the needs of healthcare providers currently using the IMCI framework, helping them to decide which patients with pneumonia not already identified for hospitalization or referral may benefit from higher-level care. As found in other studies, ^21^ this approach also underscores the high number of pneumonia patients with hypoxemia (1,421/14,509; 9.8%) that would not have been identified as requiring referral or hospitalization using the current IMCI algorithm in the absence of a pulse oximeter. In line with previous studies, greater tachypnea,^23 24 49 51^ chest indrawing,^22 23 51^ younger age,^8 22^ wheezing,^22^ respiratory distress,^8 22 24 49 51^ and low and very-low weight-for-age z-score,^8 22^ were identified as clinical signs predictive of hypoxemia in children with pneumonia. In the present study and previous studies, individual signs performed poorly in identifying hypoxemia, emphasizing the benefits of a multivariable clinical risk score.

The use of the PREPARE hypoxemia risk score and selection of cut-off to guide referral of children with suspected hypoxemia would need to be tailored to the setting of implementation. Selecting a cut-off with the highest balance between sensitivity and specificity may not be the most appropriate referral threshold in many clinical contexts.^54 55^ Care must be taken when deciding to use such a score in order to limit referral of children without hypoxemia or danger signs which could potentially come at great cost to individual patients and/or the health system. Considering that all patients with hypoxemia identified using this score would have otherwise been misidentified as not requiring referral by IMCI without pulse oximetry, using cut-offs with lower sensitivity (and higher specificity) could still make a helpful contribution, limiting unnecessary referrals, whilst still identifying many children with hypoxemia who would have been missed by the IMCI algorithm in settings where pulse oximetry is not available. Nevertheless, even utilising cut-offs of 5 or 6, where between 4-12% of all children would be referred, the majority of referrals (74-81%) would still be for children who were not hypoxemic. It may be that depending on the setting of implementation, a cut-off with relatively low specificity should consider alternative approaches to referral, such as short admission or follow-up within the community, to observe clinical evolution as proposed by Graham et al.^13^ Growing accessibility to “smartphones” might increase feasibility of a digital tool, which would allow more precise predictions and enable referral thresholds to be better tailored to the specific context of implementation. More pragmatically, elements from the PREPARE hypoxemia risk score could be combined with data on their prognostic value to predict death,^56^ to update the IMCI criteria for severe pneumonia.

The sensitivity analysis excluding respiratory distress had a higher AUROC value compared to the primary analysis including respiratory distress as a candidate predictor. This however, should not be interpreted to mean that respiratory distress does not add value in identifying hypoxemia. In fact respiratory distress had similar predictive value to identifying hypoxemia compared to lower chest indrawing and very fast breathing for age (i.e., a respiratory rate of 20 breaths or more above age cut-off values). Differences in AUROC values between our analyses and compared to other studies are influenced by differences in the underlying datasets.^52^ Indeed, this sensitivity analysis included twice the number of studies and thus the higher discrimination may reflect better generalizability of a score developed from a more diverse range of countries and settings. The value of respiratory distress should thus still be explored to improve the detection of children with hypoxemia and/or severe pneumonia within IMCI consultations, all the while considering the training, mentorship and resources required to reliably assess these signs.

There are several limitations to this study. First, we intended to derive a clinical risk score using data from primary level health facilities since this is where pulse oximetry is often inaccessible and thus the clinical risk score would be most useful. Unfortunately, due to missing data on the outcome of interest and important clinical predictors, almost all studies from primary level health facilities were excluded, leaving a cohort derived predominantly from patients presenting to hospital outpatient departments. While the exclusion of children with danger signs may reduce this spectrum bias, the high proportion of pneumonia patients with chest indrawing (62%) and hypoxemia (9.8%) compared to what is expected at primary level health facilities, demonstrates the more severe presentation of included patients. Nonetheless, a similar study that was limited to ambulatory clinics found that the sensitivity and specificity of individual variables for predicting hypoxemia were similar to hospital-based studies.^22^ Secondly, it has been reported that pulse oximetry is less accurate in patients with darker skin,^57 58^ and this was not considered in the present clinical risk score. Third, only 9% of children within the target age range with pneumonia were included (14,509/157,433). The vast majority of children and studies were excluded due to missing data (predictors or outcome). While this limited the diversity of the dataset, this decision was taken in order to prioritize better quality data, limiting bias in order to retain studies that systematically assessed the predictors and outcome of interest. Further work is needed to confirm that the score is generalizable to the whole pneumonia population more widely – assuming that patients with pulse oximetry readings are likely to be more severe, the prevalence of hypoxemia in the pneumonia population is likely lower,^59^ and the score may therefore benefit from updating.

## CONCLUSION

Hypoxemia predicts mortality at all levels of care,^2-4^ and pulse oximetry helps increase sensitivity to identify children with severe pneumonia as defined by IMCI.^6 19^ Identification of children with pneumonia who are at high risk of hypoxemia using the PREPARE hypoxemia risk score could improve recognition of children who would benefit from further management and may reduce morbidity and mortality in lower-level health facilities without access to pulse oximetry, even when other well-recognized indications of referral are absent. These benefits must be weighed against the burden of increased referrals on patients and health systems. Further validation and model updating in community settings is both a research and policy priority.

## Data Availability

Data may be made available upon reasonable request to the corresponding author.

## ACKNOWLEDGEMENTS

We would like to thank all study participants, clinicians and researchers involved in the included studies.

## AUTHOR CONTRIBUTIONS

RT, AC, FB, GL and YBN conceptualized and designed the study, interpreted the data and wrote the first draft of the manuscript. YBN verified the underlying data and conducted the statistical analyses. All authors oversaw data collection, verified the underlying data, assisted with the interpretation of the data and reviewed and provided input to the final draft. YBN had final responsibility for the decision to submit for publication.

