## Supplementary Materials for "Development and validation of a novel clinical risk score to predict hypoxemia in children with pneumonia using the WHO PREPARE dataset"

### Supplementary material

**Table S5.** Baseline characteristics of children with pneumonia (fast breathing and/or chest indrawing), pulse oximetry measurements and complete data on all candidate predictors included in the PREPARE risk score development and validation when excluding respiratory distress (n=15,592) 9

**Table S1**: Multivariable regression model for predicting SpO2<92% in the development dataset (n=10,884)

| Characteristic | SpO2 92-100% (n=9,117) | SpO2 <92% (n=1,767) | Odds ratio (95% CI) | Adjusted OR (95% CI) |
|---|---|---|---|---|
| **Age** | | | | |
| 2-5 months | 2,787 (30.6%) | 613 (34.7%) | 1.31 (1.16, 1.48) | 1.25 (1.10, 1.42) |
| 6-11 months | 2,344 (25.7%) | 486 (27.5%) | 1.24 (1.09, 1.41) | 1.31 (1.15, 1.50) |
| 12-59 months | 3,986 (43.7%) | 688 (37.8%) | (reference) | (reference) |
| **Sex** | | | | |
| Male | 5,099 (55.9%) | 1,026 (58.1%) | (reference) | (reference) |
| Female | 4,018 (44.1%) | 741 (41.9%) | 0.92 (0.83, 1.02) | 0.92 (0.83, 1.02) |
| **Pneumonia classification** | | | | |
| Fast breathing only | 3,673 (40.3%) | 379 (21.5%) | (reference) | (reference) |
| Lower chest indrawing | 5,444 (59.7%) | 1,388 (78.5%) | 2.47 (2.19, 2.79) | 1.50 (1.29, 1.73) |
| **Any sign of respiratory distress**[b] | | | | |
| No | 5,299 (58.1%) | 631 (35.7%) | (reference) | (reference) |
| Yes | 3,818 (41.9%) | 1,136 (64.3%) | 2.50 (2.25, 2.78) | 2.02 (1.78, 2.29) |
| **Respiratory rate** | | | | |
| <age-adjusted tachypnea threshold[a] | 110 (1.2%) | 14 (0.8%) | (reference) | (reference) |
| 0-9 breaths/min above cut-off | 4,117 (45.2%) | 483 (27.3%) | 0.92 (0.52, 1.62) | 1.07 (0.61, 1.90) |
| 10-19 breaths/min above cut-off | 3,298 (36.2%) | 695 (39.3%) | 1.66 (0.94, 2.90) | 1.67 (0.95, 2.94) |
| >/=20 breaths/min above cut-off | 1,592 (17.5%) | 575 (32.5%) | 2.83 (1.61, 4.99) | 2.66 (1.51, 4.69) |
| **Axillary temperature** | | | | |
| Normal (>/=35.5°C to 37.5°C) | 5,839 (64.1%) | 1,069 (60.5%) | (reference) | (reference) |
| >37.5°C | 3,173 (34.8%) | 681 (38.5%) | 1.17 (1.06, 1.30) | 1.02 (0.91, 1.14) |
| <35.5°C | 105 (1.1%) | 17 (1.0%) | 0.88 (0.53, 1.48) | 0.89 (0.52, 1.52) |
| **Weight for age z-score** | | | | |
| >=-2 | 7,427 (81.5%) | 1,350 (76.4%) | (reference) | (reference) |
| <-2 to -3 | 1,113 (12.2%) | 251 (14.2%) | 1.24 (1.07, 1.44) | 1.16 (1.00, 1.36) |
| <-3 | 577 (6.3%) | 166 (9.4%) | 1.58 (1.32, 1.90) | 1.59 (1.31, 1.93) |

[a]IMCI chartbook threshold for tachypnea: age 2-11 months >=50 breaths per minute; age 12-59 months >=40 breaths per minute; [b] head nodding or nasal flaring or grunting

**Table S2.** Components of the PREPARE SpO2 <92% risk score in the development dataset (n=10,884)

| Factor | Adjusted log coefficient | PREPARE hypoxemia risk score[a] |
|---|---|---|
| **Age** | | |
| 2-5 months | 0.22 | -- |
| 6-11 months | 0.27 | +1 |
| 12-59 months | --- | --- |
| **Pneumonia classification** | | |
| Fast breathing only | -- | --- |
| Lower chest indrawing | 0.40 | +1 |
| **Respiratory rate** | | |
| <age-adjusted tachypnea threshold[b] | -- | --- |
| 0-9 breaths/min above cut-off | 0.07 | --- |
| 10-19 breaths/min above cut-off | 0.51 | --- |
| >/=20 breaths/min above cut-off | 0.98 | +2 |
| **Any sign of respiratory distress[c]** | | |
| No | --- | -- |
| Yes | 0.70 | +1 |
| **Weight for age z-score** | | |
| >=-2 | -- | --- |
| <-2 to -3 | 0.15 | -- |
| <-3 | 0.46 | +1 |

[a]Regression coefficients of each retained parameter with a statistically significant adjusted odds ratio (confidence interval (CI) did not overlap with 1.0) was rounded to the nearest 0.5 and then doubled to derive the points contributing to the PREPARE hypoxemia risk score; [b]IMCI chartbook threshold for tachypnea: age 2-11 months >=50 breaths per minute; age 12-59 months >=40 breaths per minute; [c] head nodding or nasal flaring or grunting

**Table S3**. Performance of the PREPARE SpO2 <92% risk score at each cut-off in the validation dataset (n=3,625)

| Score | SpO2 <92%, n (%)* | Sensitivity (95% CI)* | Specificity (95 CI)* | Positive Likelihood Ratio (95% CI)* | Negative Likelihood Ratio (95% CI)* | Miss rate*† % (n/N) | False discovery rate*‡ % (n/N) | Proportion referred*^ % (n/N) |
|---|---|---|---|---|---|---|---|---|
| 0 | 37 (4.7%) | 100.0 | 0.0 | 1.00 | --- | 0% (0/275) | 85.8% (3111/3625) | 100% (3625/3625) |
| 1 | 99 (12.5%) | 92.8 (90.2, 94.9) | 24.4 (22.9, 25.9) | 1.23 (1.19, 1.27) | 0.29 (0.21, 0.40) | 7.2% (37/514) | 83.1% (2353/2830) | 78.1% (2830/3625) |
| 2 | 157 (13.6%) | 73.5 (69.5, 77.3) | 46.6 (44.8, 48.3) | 1.38 (1.29, 1.46) | 0.57 (0.49, 0.66) | 26.5% (136/514) | 81.5% (1661(2039) | 56.2% (2039/3625) |
| 3 | 105 (20.5%) | 43.0 (38.7, 47.4) | 78.8 (77.3, 80.2) | 2.02 (1.79, 2.28) | 0.72 (0.67, 0.78) | 57.0% (293/514) | 74.9% (661/882) | 24.3% (882/3625) |
| 4 | 87 (28.4%) | 22.6 (19.0, 26.4) | 91.8 (90.8, 92.8) | 2.76 (2.27, 3.37) | 0.84 (0.80, 0.88) | 77.4% (398/514) | 68.7% (255/371) | 10.2% (371/3254) |
| 5 | 26 (43.3%) | 5.6 (3.8, 8.0) | 98.8 (98.4, 99.2) | 5.01 (3.09, 8.13) | 0.95 (0.93, 0.98) | 94.4% (485/514) | 55.4% (36/65) | 1.8% (65/3625) |
| 6 | 3 (75.0%) | 0.6 (0.1, 1.7) | 100.0 (99.8, 100.0) | 18.2 (1.89, 174.0) | 0.99 (0.99, 1.00) | 99.4% (511/514) | 25% (1/4) | 0.1% (4/3625) |

*Calculated at ≥ each respective cut-off

†Miss rate (also known as false negative rate = False Negative/(True Positive + False Negative)

‡False discovery rate = False Positive/(False Positive+True Positive)

^Referral used as a proxy for identification of hypoxemia, assuming that all children identified as at risk of hypoxemia would be referred

**Table S4**: Characteristics of studies included in the development and validation of the PREPARE hypoxemia risk score without respiratory distress

| Study | Study design | Location | Age | Original inclusion criteria from the source studies | Original exclusion criteria from the source studies | Sample size included in the present analysis over the sample size from the original study, n/N (%) | Hypoxemia n/N (%) |
|---|---|---|---|---|---|---|---|
| Addo-Yobo 2004[39] | Randomized controlled trial | Colombia, Ghana, India, Mexico, Pakistan, South Africa, Vietnam, Zambia | 3-59 months | - Children with chest-indrawing pneumonia or WHO-defined severe pneumonia[a] | - Children with any danger sign during current illness (convulsions, abnormally sleepy or difficult to awake, stridor in a calm child).<br>- Children with severe comorbidities and chronic conditions: severe malnutrition, measles, HIV infection, congenital cardiac or respiratory anomalies, etc<br>- Children with other diseases requiring antibiotic therapy on presentation<br>- Children with $SpO_2$ <80%<br>- Children with known prior anaphylactic reaction to penicillin or amoxicillin<br>- Children with known antibiotic therapy for 48 hours or more prior to admission<br>- Children living outside the catchment area of the hospital<br>- children unable to receive oral medications | 1,251/1,702 (73.5%) | 13.6% (170/1,251) |
| Basnet, 2012[40] | Randomized controlled trial | Kathmandu, Nepal | 2-35 months | - Cough for <14 days<br>- Difficulty breathing ≤72 hours with presence of lower chest indrawing on examination | - Children with recurrent wheezing, heart disease,<br>other severe illness, severe malnutrition, | 192/641 (30.0%) | 55.7% (107/192) |

| | | | | | | | |
|---|---|---|---|---|---|---|---|
| | | | | | dehydration, hemoglobin <70 g/L, chronic cough, effusion on chest x-ray,<br>or history of documented tuberculosis. | | |
| Bénet, 2017[41] | Prospective case series | - Phnom Penh, Cambodia<br>- Beijing, China<br>- Port au Prince, Haiti<br>- Lucknow, India<br>- Pune-Vadu, India<br>- Antananarivo, Madagascar<br>- Bamako, Mali<br>- Ulaanbaatar, Mongolia<br>- San Lorenzo, Paraguay. | 2-59 months | - Children with WHO-defined pneumonia with first symptoms<br>lasting <14 days<br>- Children with radiographic pneumonia (WHO criteria) | - Children with wheezing at auscultation | 106/888 (11.9%) | 5.7% (6/106) |
| Clara WS. Unpublished data 2012[42] | Retrospective cohort | Chariqui Province, Panama | 2-59 months | - Children with WHO-defined pneumonia of any severity[b] | None | 9/85 (10.6%) | 44.4% (4/9) |
| Cutts, 2005[38] | Randomized controlled trial | Eastern Gambia | 40-364 days | Children with WHO-defined pneumonia of any severity[b] | - Intent to move out within 4 months<br>- Previous or uncertain receipt of diphtheria-pertussis-tetanus/*Haemophilus influenzae* type b (DPT/Hib) or DPT vaccine<br>- Inclusion in a previous vaccine trial<br>- Serious chronic illness | 323/1,716 (18.8%) | 4.3% (14/323) |
| Ferolla, 2013[43] | Prospective cohort | Buenos Aires, Argentina | < 2 years | - Children <2 years admitted with a diagnosis of severe acute respiratory illness, defined as the sudden onset of cough, wheezing, retractions and/or crackles with or without fever | None | 59/5,043 (1.2%) | 39.0% (23/59) |
| Gessner, 2005[44] | Prospective cohort | Lombok, Indonesia | 2-35 months | - Children with WHO-defined pneumonia of any severity[b] | - Received three doses of diphtheria-tetanus-<br>pertussis (DTP) vaccine | 4,464/6,221 (71.8%) | 34.5% (1,542/4,464 |
| Klugman, 2003[45] | Randomized controlled trial | Johannesburg, South Africa | 2-59 months | ICD-10<br>codes for pneumonia | - Children with progressive underlying neurological disorder, history of seizures, or infantile spasms | 1,517/10,114 (15.0%) | 16.9% (256/1,517) |

| | | | | | | | |
|---|---|---|---|---|---|---|---|
| | | | | | - Low likelihood of receiving three doses of vaccine | | |
| Lazzerini, 2016[46] | Prospective cohort | Malawi | 2-59 months | - Children with WHO-defined pneumonia of any severity[b] | None | 2,640/16,475 (16.0%) | 4.8% (127/2,640) |
| Mathew, 2015[47] | Prospective cohort | Chandigarh, India | 2-59 months | - Children with WHO-defined pneumonia of any severity[b] | - Duration of illness >7 days<br>- Antibiotics for >24 hours<br>- Previous hospitalization within the preceding 30 days<br>- Children with wheeze who received a single dose of bronchodilator and whose symptoms disappeared | 510/2400 (21.3%) | 10.2% (52/510) |
| McCollum, 2017[48] | Prospective cohort | Mchinji and Lilongwe Districts, Malawi | 2-59 months | - Children with WHO-defined pneumonia of any severity[b] | None | 3,601/6,764 (53.2%) | 3.6% (129/3,601) |
| O'Grady, 2012[49] | Randomized controlled trial | Central Australia | 2-59 months | - Children with WHO-defined pneumonia of any severity[b] | Children with wheezing and chronic conditions | 26/147 (17.7%) | 0.0% (0/26) |
| Ugpo, 2009[50] | Randomized controlled trial | Bohol, Philippines | 2-59 months | - Children with WHO-defined pneumonia of any severity[b] | - Children with known seizure disorder presenting with febrile convulsions<br>- Children with a history of benign febrile convulsion who present with fever and seizure with normal neurologic examination<br>- Children with dengue shock syndrome | 586/1,201 (48.8%) | 14.0% (82/586) |
| Wadhwa, 2013[51] | Randomized controlled trial | New Delhi, India | 2–24 months | - Children with WHO-defined pneumonia of any severity[b]<br>- Crepitations on auscultation | - Need for mechanical ventilation or inotropic medications<br>- Any other serious underlying medical condition | 308/550 (56.0%) | 1.0% (3/308) |

[a]WHO-defined severe pneumonia: presence of danger signs, stridor, central cyanosis, severe respiratory distress (nasal flaring, grunting, head-nodding), in children with cough and/or difficulty breathing

[b]WHO-defined pneumonia of any severity: presence of age-specific fast breathing, lower chest indrawing, or danger signs, in children with a cough and/or difficulty breathing.

**Table S5.** Baseline characteristics of children with pneumonia (fast breathing and/or chest indrawing), pulse oximetry measurements and complete data on all candidate predictors included in the PREPARE risk score development and validation when excluding respiratory distress (n=15,592)

| Characteristic | Total (n=15,592) | Development dataset (n=11,731) | Validation dataset (n=3,861) |
|---|---|---|---|
| **Age**[a] | | | |
| 2-5 months | 5,147 (33.0%) | 4,041 (34.5%) | 1,106 (28.6%) |
| 6-11 months | 4,257 (27.3%) | 3,313 (28.2%) | 944 (25.5%) |
| 12-59 months | 6,188 (39.7%) | 4,377 (37.3%) | 1,811 (46.9%) |
| **Sex** | | | |
| Male | 8,022 (51.5%) | 6,015 (51.3%) | 2,007 (52.0%) |
| Female | 7,570 (48.5%) | 5,716 (48.7%) | 1,854 (48.0%) |
| **Pneumonia classification** | | | |
| Fast breathing only | 6,061 (38.9%) | 4,489 (38.3%) | 1,572 (40.7%) |
| Lower chest indrawing | 9,531 (61.1%) | 7,242 (61.7%) | 2,289 (59.3%) |
| **Respiratory rate** | | | |
| <age-adjusted tachypnea threshold[b] | 204 (1.3%) | 90 (0.8%) | 114 (3.0%) |
| 0-9 breaths/min above cut-off | 6,657 (42.7%) | 4,995 (42.6%) | 1,662 (43.1%) |
| 10-19 breaths/min above cut-off | 5,758 (36.9%) | 4,351 (37.1%) | 1,407 (36.4%) |
| >/=20 breaths/min above cut-off | 2,973 (19.1%) | 2,295 (19.6%) | 678 (17.5%) |
| **Body temperature** | | | |
| Normal (>/=35.5°C to 37.5°C) | 10,252 (65.7%) | 7,678 (65.4%) | 2,574 (66.7%) |
| >=37.5°C | 5,250 (33.7%) | 3,986 (34.0%) | 1,264 (32.7%) |
| <35.5°C | 90 (0.6%) | 67 (0.6%) | 23 (0.6%) |
| **Weight for age z-score** | | | |
| >=-2 | 11,690 (75.0%) | 8,716 (74.3%) | 2,974 (77.0%) |
| <-2 to -3 | 2,293 (14.7%) | 1,770 (15.1%) | 523 (13.6%) |
| <-3 | 1,609 (10.3%) | 1,245 (10.6%) | 364 (9.4%) |
| **Oxygen saturation ($SpO_2$)** | | | |
| >=90% | 13,077 (83.9%) | 9,767 (83.3%) | 3,310 (85.7%) |
| <90% | 2,515 (16.1%) | 1,964 (16.7%) | 551 (14.3%) |

[a]Some studies did not include patients in the whole 2-59 month age range; [b]IMCI chartbook threshold for tachypnea: age 2-11 months >=50 breaths per minute; age 12-59 months >=40 breaths per minute

**Table S6:** Multivariable regression model for predicting hypoxemia in the development dataset without respiratory distress as a candidat e predictor (n=11,731)

| Characteristic | No hypoxemia (n=9,767) | Hypoxemia (n=1,964) | Odds ratio (95% CI) | Adjusted OR (95% CI) |
|---|---|---|---|---|
| **Age** | | | | |
| 2-5 months | 3,178 (32.5%) | 863 (43.9%) | 2.13 (1.89, 2.04) | 1.92 (1.69, 2.19) |
| 6-11 months | 2,706 (27.7%) | 607 (30.9%) | 1.76 (1.55, 2.00) | 1.65 (1.44, 1.90) |
| 12-59 months | 3,883 (39.8%) | 4947 (25.2%) | (reference) | (reference) |
| **Sex** | | | | |
| Male | 5,083 (52.0%) | 932 (47.4%) | (reference) | (reference) |
| Female | 4,684 (48.0%) | 1,032 (52.6%) | 1.20 (1.09, 1.32) | 1.25 (1.12, 1.39) |
| **Pneumonia classification** | | | | |
| Fast breathing only | 4,257 (43.6%) | 232 (11.8%) | (reference) | (reference) |
| Lower chest indrawing | 5,510 (56.4%) | 1,732 (88.2%) | 5.77 (5.00, 6.65) | 4.44 (3.83, 5.15) |
| **Respiratory rate** | | | | |
| <age-adjusted tachypnea threshold* | 79 (0.8%) | 11 (0.6%) | (reference) | (reference) |
| 0-9 breaths/min above cut-off | 4,475 (45.8%) | 520 (26.5%) | 0.83 (0.4, 1.58) | 0.90 (0.46, 1.77) |
| 10-19 breaths/min above cut-off | 3,500 (35.8%) | 851 (43.3%) | 1.75 (0.93, 3.30) | 1.44 (0.74, 2.81) |
| >/=20 breaths/min above cut-off | 1,713 (17.5%) | 582 (29.6%) | 2.44 (1.29, 4.62) | 2.10 (1.08, 4.10) |
| **Body temperature** | | | | |
| Normal (>/=35.5°C to 37.5°C) | 6,476 (66.3%) | 1,202 (61.2%) | (reference) | (reference) |
| >37.5°C | 3,230 (33.1%) | 756 (38.5%) | 1.26 (1.14, 1.39), | 1.17 (1.05, 1.31) |
| <35.5°C | 61 (0.6%) | 6 (0.3%) | 0.53 (0.23, 1.23) | 0.75 (0.33, 1.73) |
| **Weight for age z-score** | | | | |
| >=-2 | 7,494 (76.7%) | 1,222 (62.2%) | (reference) | (reference) |
| <-2 to -3 | 1,367 (14.0%) | 403 (20.5%) | 1.81 (1.59, 2.05)2.29 (2.00, 2.64) | 1.67 (1.46, 1.91) |
| <-3 | 906 (9.3%) | 339 (17.3%) | | 1.93 (1.66, 2.25) |

*IMCI chartbook threshold for tachypnea: age 2-11 months >=50 breaths per minute; age 12-59 months >=40 breaths per minute

**Table S7.** Components of the PREPARE hypoxemia risk score in the development dataset without respiratory distress as a candidate predictor (n=11,731)

| Factor | Adjusted log coefficient | PREPARE hypoxemia risk score* |
|---|---|---|
| **Age** | | |
| 2-5 months | 0.65 | +1 |
| 6-11 months | 0.50 | +1 |
| 12-59 months | -- | --- |
| **Pneumonia classification** | | |
| Fast breathing only | -- | --- |
| Lower chest indrawing | 1.49 | +3 |
| **Respiratory rate** | | |
| <threshold | -- | --- |
| 0-9 breaths above cut-off | -0.10 | --- |
| 10-19 breaths above cut-off | 0.37 | --- |
| >/=20 breaths above cut-off | 0.74 | +1 |
| **Weight for age z-score** | | |
| >=-2 | -- | --- |
| <-2 to -3 | 0.51 | +1 |
| <-3 | 0.66 | +1 |

**Table S8**. Performance of the PREPARE hypoxemia risk score without respiratory distress at each cut-off in the validation dataset (n=3,861)

| Score | Hypoxemia, n (%) | Sensitivity (95% CI)‡^* ^ | Specificity (95 CI)‡ * ^ | Positive Likelihood Ratio (95% CI)‡ | Negative Likelihood Ratio (95% CI)‡ | Miss rate† % (n/N) | False discovery rate‡ % (n/N) | Proportion referred^ % (n/N) |
|---|---|---|---|---|---|---|---|---|
| 0 | 3 (0.5%) | 100.0 | 0.0 | 1.00 | --- | 0% (0/551) | 85.7% (3310/3861) | 100% (3861/3861) |
| 1 | 24 (3.1%) | 99.5 (98.4, 99.9) | 19.6 (18.3, 21.0) | 1.24 (1.22, 1.26) | 0.28 (0.01, 0.09) | 0.5% (3/551) | 82.9% (2660/3208) | 83.1% (3208/3861) |
| 2 | 6 (4.5%) | 95.1 (93.0, 96.7) | 42.5 (40.8, 44.2) | 1.65 (1.60, 1.71) | 0.11 (0.08, 0.17) | 4.9% (27/551) | 78.4% (1904/2428) | 62.9% (2428/3861) |
| 3 | 46 (11.0%) | 94.0 (91.7, 95.8) | 46.3 (44.6, 48.0) | 1.75 (1.69, 1.82) | 0.13 (0.09, 0.18) | 6.0% (33/551) | 77.4% (1777/2295) | 59.4% (2295/3861) |
| 4 | 278 (22.2%) | 85.7 (82.5, 88.5) | 57.6 (55.9, 59.3) | 2.02 (1.92, 2.13) | 0.25 (0.20, 0.31) | 14.3% (79/551) | 74.8% (1403/1875) | 48.6% (1875/3861) |
| 5 | 170 (30.1%) | 35.2 (31.2, 39.4) | 87.1 (85.9, 88.3) | 2.74 (2.37, 3.16) | 0.74 (0.70, 0.79) | 64.8% (357/551) | 68.7% (426/620) | 16.1% (620/3861) |
| 6 | 24 (42.9%) | 4.4 (2.8, 6.4) | 99.0 (98.6, 99.3) | 4.51 (2.67, 7.59) | 0.97 (0.95, 0.98) | 95.6% (527/551) | 57.1% (32/56) | 1.5% (56/3861) |

*Calculated at ≥ each respective cut-off

†Miss rate (also known as false negative rate = False Negative/(True Positive + False Negative)

‡False discovery rate = False Positive/(False Positive+True Positive)

^Referral used as a proxy for identification of hypoxemia, assuming that all children identified as at risk of hypoxemia would be referred

**Table S9:** Characteristics of studies included in the development and validation of the PREPARE hypoxemia risk score when including wheezing as a candidate predictor

| Study | Study design | Location | Age | Original inclusion criteria from the source studies | Original exclusion criteria from the source studies | Sample size included in the present analysis over the sample size from the original study, n/N (%) | Hypoxemia % (n/N) |
|---|---|---|---|---|---|---|---|
| Basnet, 2012[29] | Randomized controlled trial | Kathmandu, Nepal | 2-35 months | - Cough for <14 days<br>- Difficulty breathing ≤72 hours with presence of lower chest indrawing on examination | - Children with recurrent wheezing, heart disease, other severe illness, severe malnutrition, dehydration, hemoglobin <70 g/L, chronic cough, effusion on chest x-ray, or history of documented tuberculosis. | 378/641 (59.0%) | 66.4% (251/378) |
| Cutts, 2005[32] | Randomized controlled trial | Eastern Gambia | 40-364 days | Children with WHO-defined pneumonia of any severity* | - Intent to move out within 4 months<br>- Previous or uncertain receipt of diphtheria-pertussis-tetanus/*Haemophilus influenzae* type b (DPT/Hib) or DPT vaccine<br>- Inclusion in a previous vaccine trial<br>- Serious chronic illness | 433/1,716 (25.2%) | 5.1% (22/433) |
| Lazzerini, 2016[36] | Prospective cohort | Malawi | 2-59 months | - Children with WHO-defined pneumonia of any severity* | None | 7254/16,475 (44.0%) | 9.5% (687/7254) |
| Mathew, 2015[37] | Prospective cohort | Chandigarh, India | 2-59 months | - Children with WHO-defined pneumonia of any severity* | - Duration of illness >7 days<br>- Antibiotics for >24 hours<br>- Previous hospitalization within the preceding 30 days | 1132/2400 (47.2%) | 11.8% (134/1132) |

| | | | | | | | |
|---|---|---|---|---|---|---|---|
| | | | | | - Children with wheeze who received a single dose of bronchodilator and whose symptoms disappeared | | |
| McCollum, 2017[38] | Prospective cohort | Mchinji and Lilongwe Districts, Malawi | 2-59 months | - Children with WHO-defined pneumonia of any severity* | None | 4253/6,764 (62.9%) | 6.2% (265/4253) |
| O'Grady, 2012[39] | Randomized controlled trial | Central Australia | 2-59 months | - Children with WHO-defined pneumonia of any severity* | None | 26/147 (17.7%) | 0.0% (0/26) |
| Wadhwa, 2013[41] | Randomized controlled trial | New Delhi, India | 2–24 months | - Children with WHO-defined pneumonia of any severity*<br>- Crepitations on auscultation | - Need for mechanical ventilation or inotropic medications<br>- Any other serious underlying medical condition | 316/550 (56.0%) | 0.9% (3/316) |

*WHO-defined pneumonia of any severity: presence of age-specific fast breathing, lower chest indrawing, or danger signs or stridor in a calm child, in children with a cough or difficulty breathing.

**Table S10.** Baseline characteristics of children with pneumonia (fast breathing and/or chest indrawing), pulse oximetry measurements and complete data on all candidate predictors included in the PREPARE risk score development and validation when including wheezing as a candidate predictor (n=13,792)

| Characteristic | Total (n=13,792) | Development dataset (n=10,397) | Validation dataset (n=3,395) |
|---|---|---|---|
| **Age**[a] | | | |
| 2-5 months | 4,050 (29.4%) | 3,220 (31.0%) | 830 (24.4%) |
| 6-11 months | 3,475 (25.2%) | 2,705 (26.0%) | 770 (22.7%) |
| 12-59 months | 6,267 (45.4%) | 4,472 (43.0%) | 1,795 (52.9%) |
| **Sex** | | | |
| Male | 7,767 (56.3%) | 5,843 (56.2%) | 1,924 (56.7%) |
| Female | 6,025 (43.7%) | 4,554 (43.8%) | 1,471 (43.3%) |
| **Pneumonia classification** | | | |
| Fast breathing only | 5,416 (39.3%) | 3,977 (38.2%) | 1,439 (42.4%) |
| Lower chest indrawing | 8,376 (60.7%) | 6,420 (61.8%) | 1,956 (57.6%) |
| **Respiratory rate** | | | |
| <age-adjusted tachypnea threshold[b] | 157 (1.1%) | 120 (1.1%) | 37 (1.1%) |
| 0-9 breaths/min above cut-off | 5,881 (42.6%) | 4,428 (42.6%) | 1,453 (42.8%) |
| 10-19 breaths/min above cut-off | 5,047 (36.6%) | 3,790 (36.4%) | 1,257 (37.0%) |
| >/=20 breaths/min above cut-off | 2,707 (19.6%) | 2,059 (19.8%) | 648 (19.1%) |
| **Wheeze** | | | |
| No | 9,805 (71.1%) | 7,496 (72.1%) | 2,309 (68.0%) |
| Yes | 3,987 (28.9%) | 2,901 (27.9%) | 1,086 (32.0%) |
| **Any sign of respiratory distress**[c] | | | |
| No | 7,791 (56.5%) | 5,823 (56.0%) | 1,968 (58.0%) |
| Yes | 6,001 (43.5%) | 4,574 (44.0%) | 1,427 (42.0%) |
| **Axillary temperature** | | | |
| Normal (>/=35.5°C to 37.5°C) | 8,940 (64.8%) | 6,619 (63.7%) | 2,321 (68.4%) |
| >=37.5°C | 4,708 (34.2%) | 3,663 (35.2%) | 1,045 (30.8%) |
| <35.5°C | 144 (1.0%) | 115 (1.1%) | 29 (0.9%) |
| **Weight for age z-score** | | | |
| >=-2 | 11,155 (80.9%) | 8,364 (80.5%) | 2,791 (82.2%) |
| <-2 to -3 | 1,709 (12.4%) | 1,316 (12.7%) | 393 (11.6%) |
| <-3 | 928 (6.7%) | 717 (6.9%) | 211 (6.2%) |
| **Oxygen saturation ($SpO_2$)** | | | |
| >=90% | 12,430 (90.1%) | 9,296 (89.4%) | 3,134 (92.3%) |
| <90% | 1,362 (9.9%) | 1,101 (10.6%) | 261 (7.7%) |

[a]Some studies did not include patients in the whole 2-59 month age range; [b]IMCI chartbook threshold for tachypnea: age 2-11 months >=50 breaths per minute; age 12-59 months >=40 breaths per minute; [c] head nodding or nasal flaring or grunting

**Table S11.** Multivariable regression model for predicting hypoxemia in the development dataset when including wheezing as a candidate predictor (n=13,792)

| Characteristic | No hypoxemia (n=9,296) | Hypoxemia (n=1,101) | Odds ratio (95% CI) | Adjusted OR (95% CI) |
|---|---|---|---|---|
| **Age** | | | | |
| 2-5 months | 2,833 (30.5%) | 387 (35.2%) | 1.38 (1.19, 1.60) | 1.34 (1.14, 1.56) |
| 6-11 months | 2,393 (25.7%) | 312 (28.3%) | 1.32 (1.13, 1.54) | 1.44 (1.22, 1.70) |
| 12-59 months | 4,070 (43.8%) | 402 (36.5%) | (reference) | (reference) |
| **Sex** | | | | |
| Male | 5,196 (55.9%) | 647 (58.8%) | (reference) | (reference) |
| Female | 4,100 (44.1%) | 454 (41.2%) | 0.89 (0.78, 1.01) | 0.90 (0.79, 1.03) |
| **Pneumonia classification** | | | | |
| Fast breathing only | 3,764 (40.5%) | 213 (19.3%) | (reference) | (reference) |
| Lower chest indrawing | 5,532 (59.5%) | 888 (80.7%) | 2.84 (2.43, 3.31) | 1.60 (1.34, 1.93) |
| **Respiratory rate** | | | | |
| <age-adjusted tachypnea threshold[a] | 110 (1.2%) | 10 (0.9%) | (reference) | (reference) |
| 0-9 breaths/min above cut-off | 4,155 (44.7%) | 273 (24.8%) | 0.72 (0.37, 1.40) | 0.87 (0.45, 1.70) |
| 10-19 breaths/min above cut-off | 3,371 (36.3%) | 419 (38.1%) | 1.37 (0.71, 2.63) | 1.39 (0.72, 2.69) |
| >/=20 breaths/min above cut-off | 1,660 (17.9%) | 399 (36.2%) | 2.64 (1.37, 5.10) | 2.47 (1.27, 4.79) |
| **Wheeze** | | | | |
| No | 6,809 (73.2%) | 687 (62.4%) | (reference) | (reference) |
| Yes | 2,487 (26.8%) | 414 (37.6%) | 1.65 (1.45, 1.88) | 1.28 (1.11, 1.47) |
| **Any sign of respiratory distress[b]** | | | | |
| No | 5,453 (58.7%) | 370 (33.6%) | (reference) | (reference) |
| Yes | 3,843 (41.3%) | 731 (66.4%) | 2.80 (2.46, 3.20) | 2.25 (1.84, 2.52) |
| **Body temperature** | | | | |
| Normal (>/=35.5°C to 37.5°C) | 5,964 (64.2%) | 655 (59.5%) | (reference) | (reference) |
| >37.5°C | 3,227 (34.7%) | 436 (39.6%) | 1.23 (1.08, 1.40) | 1.06 (0.93, 1.22) |
| <35.5°C | 105 (1.1%) | 10 (0.9%) | 0.87 (0.45, 1.67) | 0.94 (0.48, 1.85) |
| **Weight for age z-score** | | | | |
| >=-2 | 7,562 (81.3%) | 802 (72.8%) | (reference) | (reference) |
| <-2 to -3 | 1,141 (12.3%) | 175 (15.9%) | 1.45 (1.21, 1.72) | 1.35 (1.12, 1.62) |
| <-3 | 593 (6.4%) | 124 (11.3%) | 1.97 (1.60, 2.42) | 1.98 (1.59, 2.48) |

[a] IMCI chartbook threshold for tachypnea: age 2-11 months >=50 breaths per minute; age 12-59 months >=40 breaths per minute; [b] head nodding or nasal flaring or grunting

**Table S12.** Components of the PREPARE hypoxemia risk score in the development dataset when including wheezing as a candidate predictor (n=10,397)

| Factor | Adjusted log coefficient | PREPARE hypoxemia risk score including wheezing* |
|---|---|---|
| **Age** | | |
| 2-5 months | 0.29 | +1 |
| 6-11 months | 0.37 | +1 |
| 12-59 months | --- | -- |
| **Pneumonia classification** | | |
| Fast breathing only | -- | -- |
| Lower chest indrawing | 0.47 | +1 |
| **Wheeze** | | |
| No | --- | -- |
| Yes | 0.24 | +0 |
| **Respiratory rate** | | |
| < age-adjusted tachypnea threshold[a] | -- | -- |
| 0-9 breaths/min above cut-off | -0.14 | -- |
| 10-19 breaths/min above cut-off | 0.33 | -- |
| >/=20 breaths/min above cut-off | 0.90 | +2 |
| **Any sign of respiratory distress[b]** | | |
| No | -- | -- |
| Yes | 0.77 | +2 |
| **Weight for age z-score** | | |
| >=-2 | -- | +0 |
| -2 to -3 | 0.30 | +1 |
| <-3 | 0.68 | +1 |

*Regression coefficients of each retained parameter with a statistically significant adjusted odds ratio (confidence interval (CI) did not overlap with 1.0) was rounded to the nearest 0.5 and then doubled to derive the points contributing to the PREPARE hypoxemia risk score. Maximum score is 7.

[a]IMCI chartbook threshold for tachypnea: age 2-11 months >=50 breaths per minute; age 12-59 months >=40 breaths per minute; [b] head nodding or nasal flaring or grunting

**Table S13.** Comparison of risk score weights from the main analysis and the sensitivity analyses

| | Main score | < 92% | Without respiratory distress | With wheezing |
|---|---|---|---|---|
| **Age 2-5 months** | 1 | - | 1 | 1 |
| **Age 6-11 monts** | 1 | 1 | 1 | 1 |
| **Lower chest indrawing** | 1 | 1 | 3 | 1 |
| **Sign of respiratory distress** | 2 | 1 | - | 2 |
| **Respiratory rate >/= 20 breaths/min[a]** | 2 | 2 | 1 | 2 |
| **WAZ <-2 to 3** | 1 | - | 1 | 1 |
| **WAZ <-3** | 1 | 1 | 1 | 1 |
| **TOTAL** | **7** | **6** | **6** | **7** |

WAZ: Weight-for-age z-score

[a] above age-adjusted tachypnea threshold. IMCI chartbook threshold for tachypnea: age 2-11 months >=50 breaths per minute; age 12-59 months >=40 breaths per minute

**Figure S1.** Decision curve analysis of the PREPARE hypoxemia model

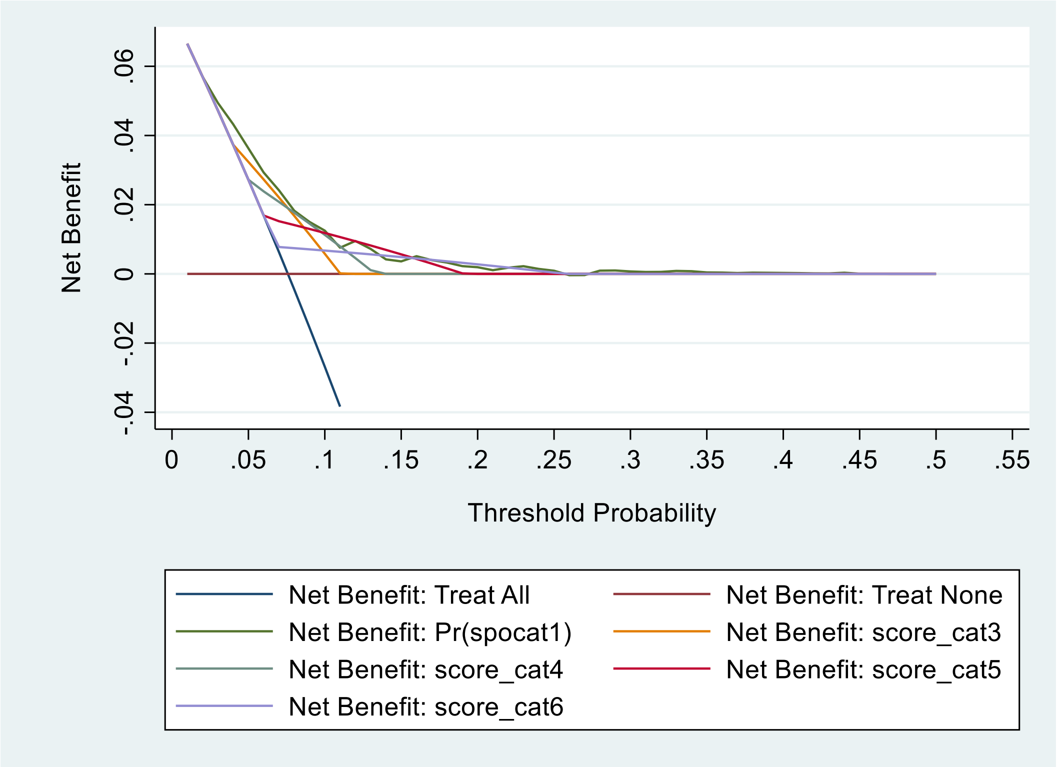

**Legend:** The net benefit of the different score thresholds (score_cat4 = refer children with a score of >/= 4; score_cat5 = refer children with a score of >/= 5; score_cat6 = refer children with a score of >/= 6; and Pr(spocat1) = Children with a predicted probability of hypoxemia greater than or equal to the threshold probability [referral threshold] are referred) are compared to a "Treat all" approach where all children are referred, and a "Treat None" approach where no children are referred. A threshold probability of 20% indicates a management strategy whereby any child with a ≥ 20% probability of being hypoxemic is referred (i.e., a scenario where the value of one correct referral is equivalent to four incorrect referrals or a number-needed-to-refer [NNR] of 5).

**Figure S2.** Receiver operating characteristic curve for the PREPARE SpO2<92% clinical prediction model for children 2–59 months of age with pneumonia

**A. Development dataset (n=10,884), AUROC 0.67, 95% CI 0.67 to 0.69**

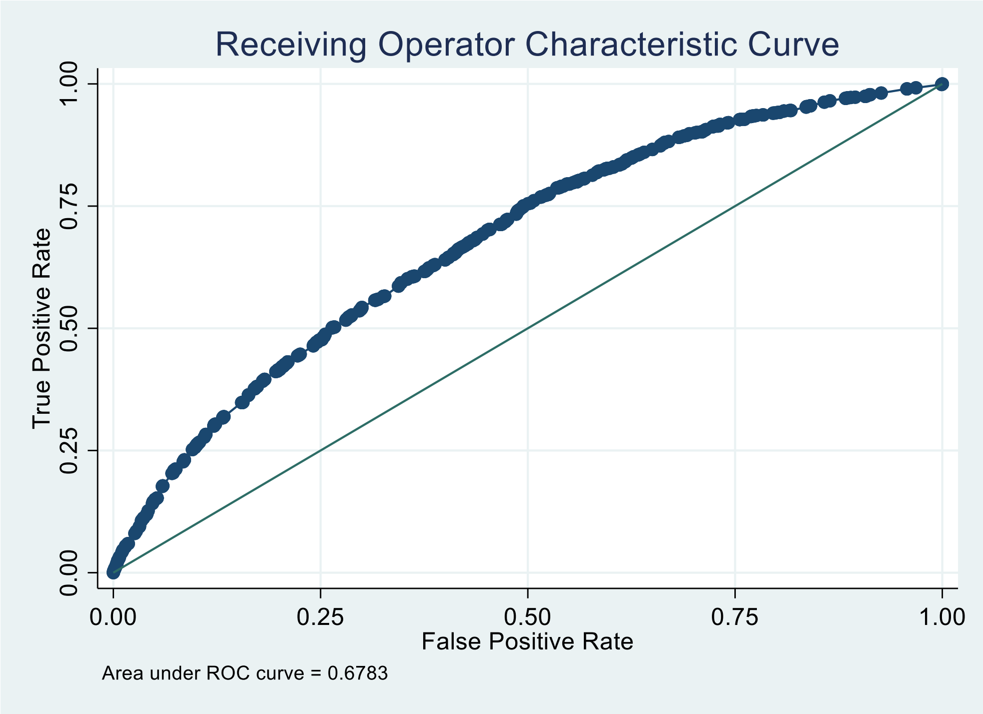

**B. Validation dataset (n=3,625), AUROC 0.67, 95% CI 0.65 to 0.70**

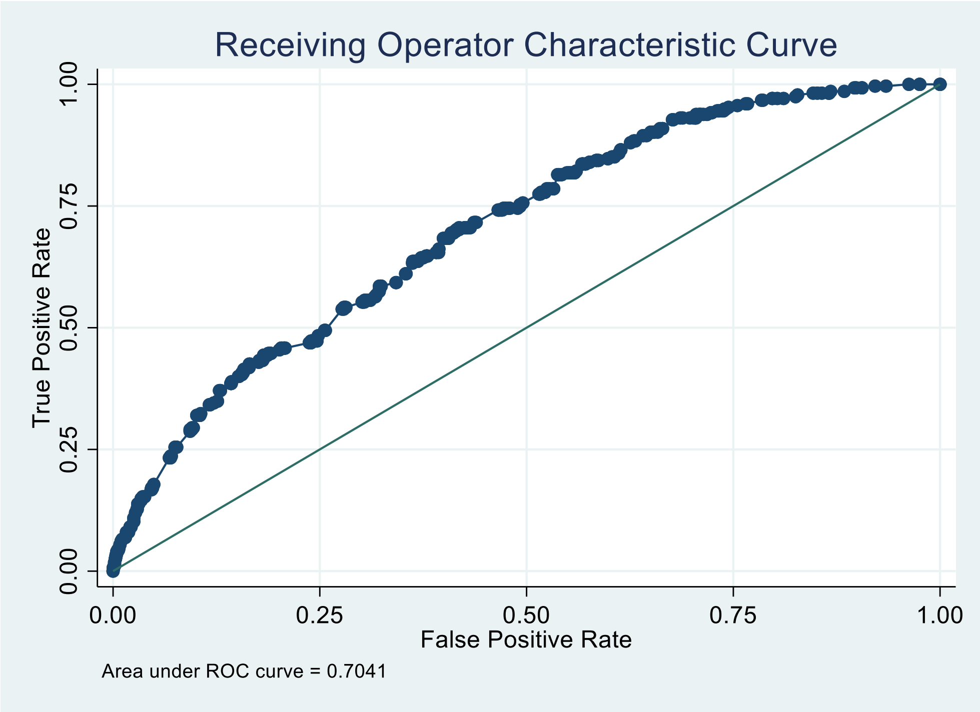

**Figure S3.** Calibration plot of the PREPARE $SpO_2$ <92% clinical prediction model in the validation dataset

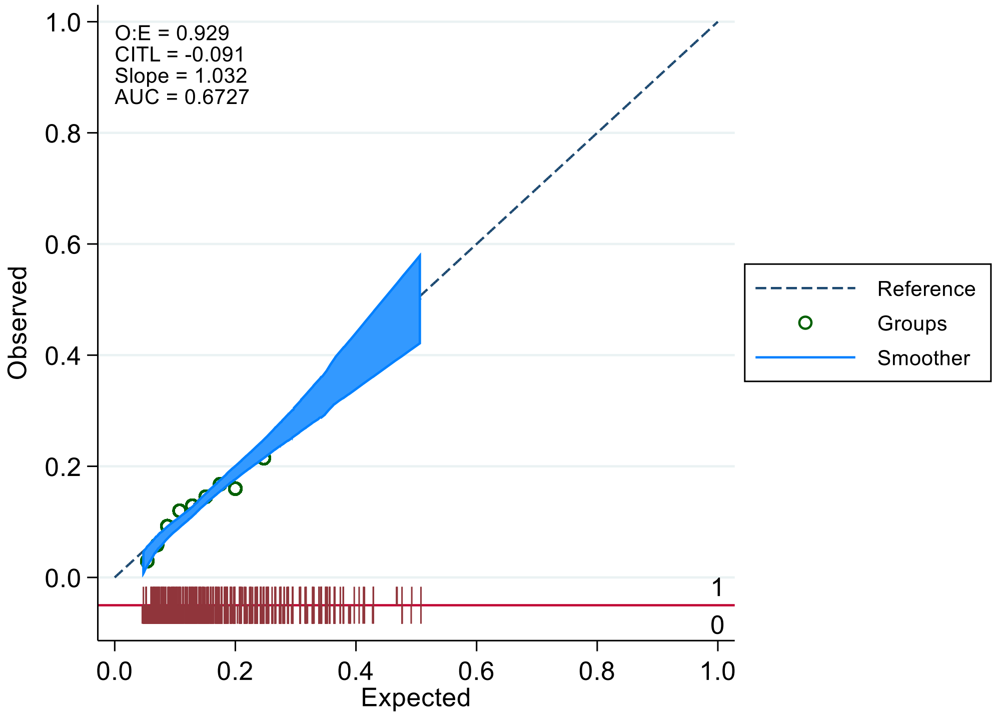

**Legend:** Reference (dashed line) indicates perfect calibration; Lowess (blue line) indicates calibration slope; Red rug plots indicate the distribution of predicted risk of participants with hypoxemia (above the line), and those without hypoxemia (below the line). E:O = expected to observed events ratio; CITL = calibration-in-the-large (intercept); AUC = area under the receiver operating characteristic curve.

**Figure S4** Receiver operating characteristic curve for the PREPARE hypoxemia clinical prediction model excluding respiratory distress as a candidate predictor for children 2–59 months of age with pneumonia

**A. Development dataset (n=11,731), AUROC 0.73, 95% CI 0.72 to 0.75**

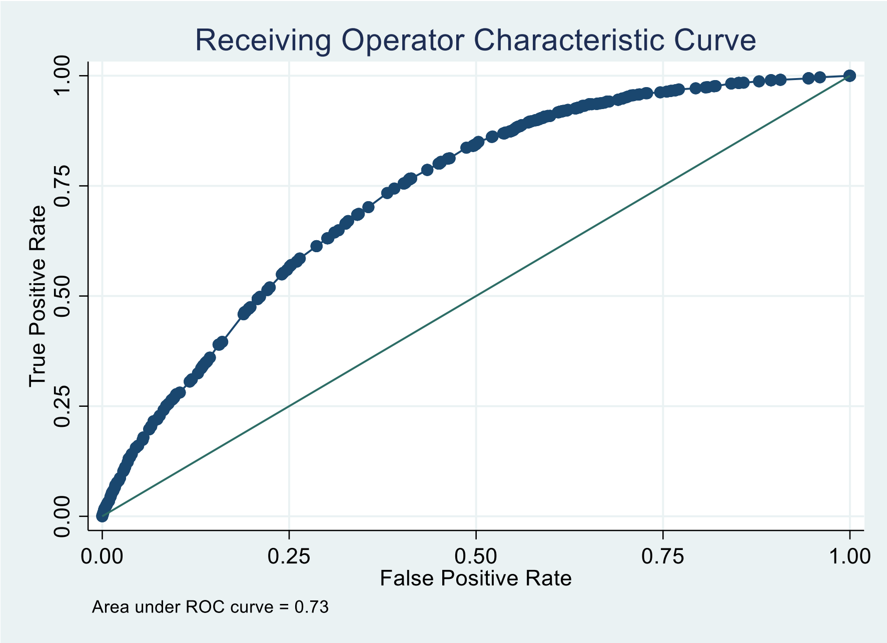

**B. Validation dataset (n=3,861), AUROC 0.78, 95% CI 0.76 to 0.80**

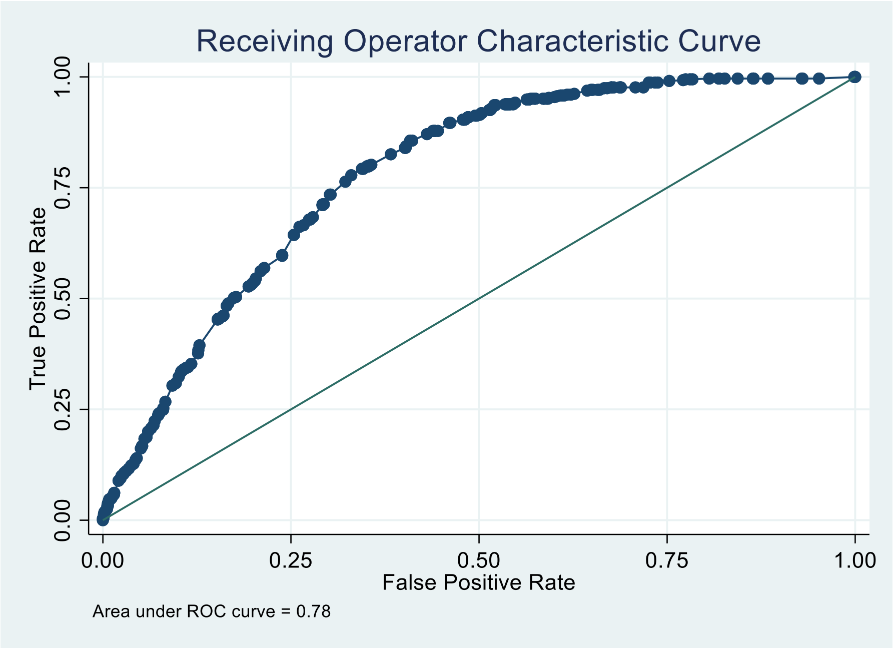

**Figure S5**: Calibration plot of the PREPARE hypoxemia clinical prediction model in the validation dataset when excluding respiratory distress as a candidate predictor

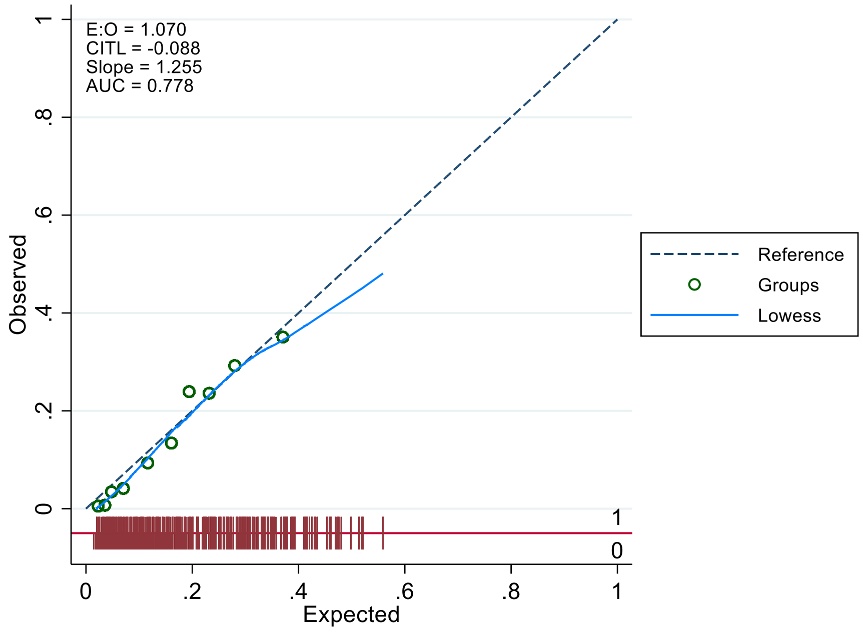

**Legend:** Reference (dashed line) indicates perfect calibration; Lowess (blue line) indicates calibration slope; Red rug plots indicate the distribution of predicted risk of participants with hypoxemia (above the line), and those without hypoxemia (below the line). E:O = expected to observed events ratio; CITL = calibration-in-the-large (intercept); AUC = area under the receiver operating characteristic curve.

**Figure S6**. Receiver operating characteristic curve for the PREPARE hypoxemia clinical prediction model when including wheezing as a candidate predictor for children 2–59 months of age with pneumonia

**A. Development dataset (n=10,397), AUROC 0.71, 95% CI 0.69 to 0.72**

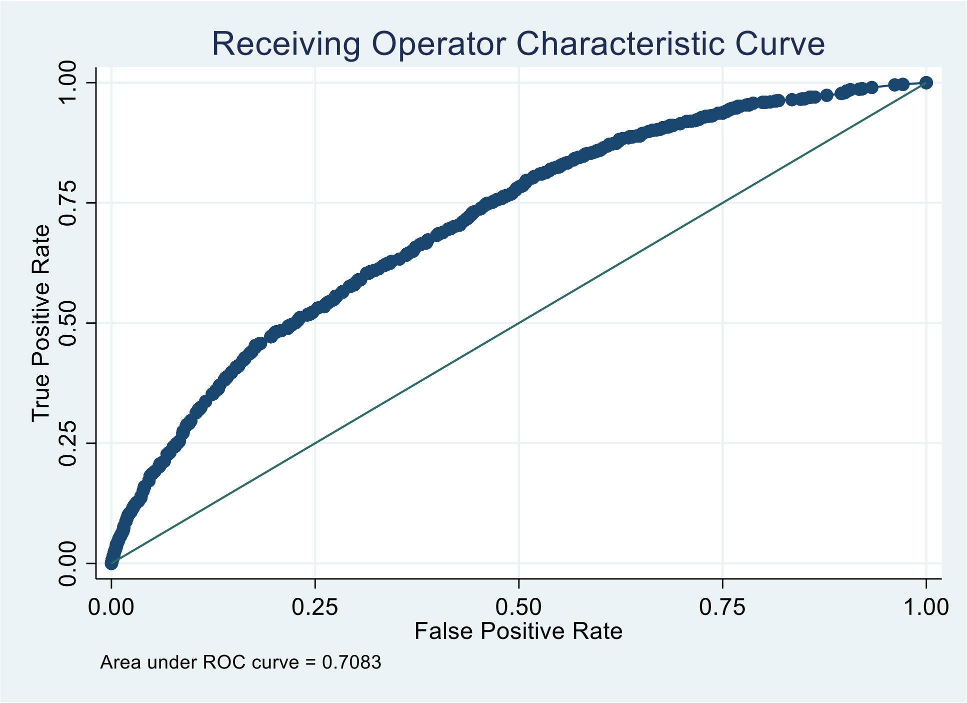

**B. Validation dataset (n=3,395), AUROC 0.72, 95% CI 0.69 to 0.75**

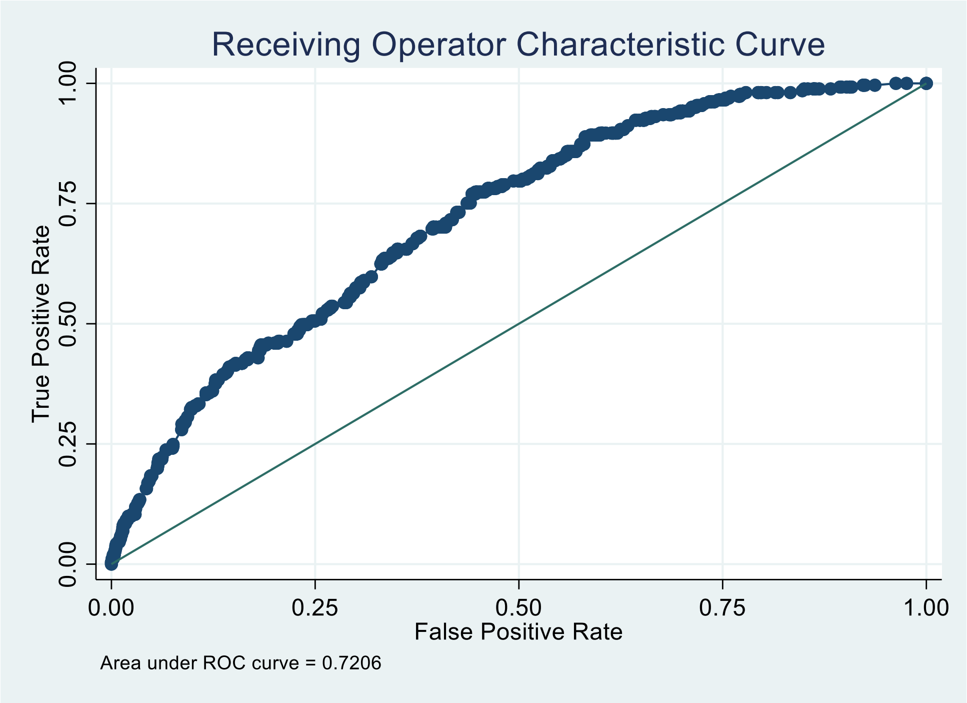

**Figure S7**: Calibration plot of the PREPARE hypoxemia clinical prediction model in the validation dataset when including wheezing as a candidate predictor

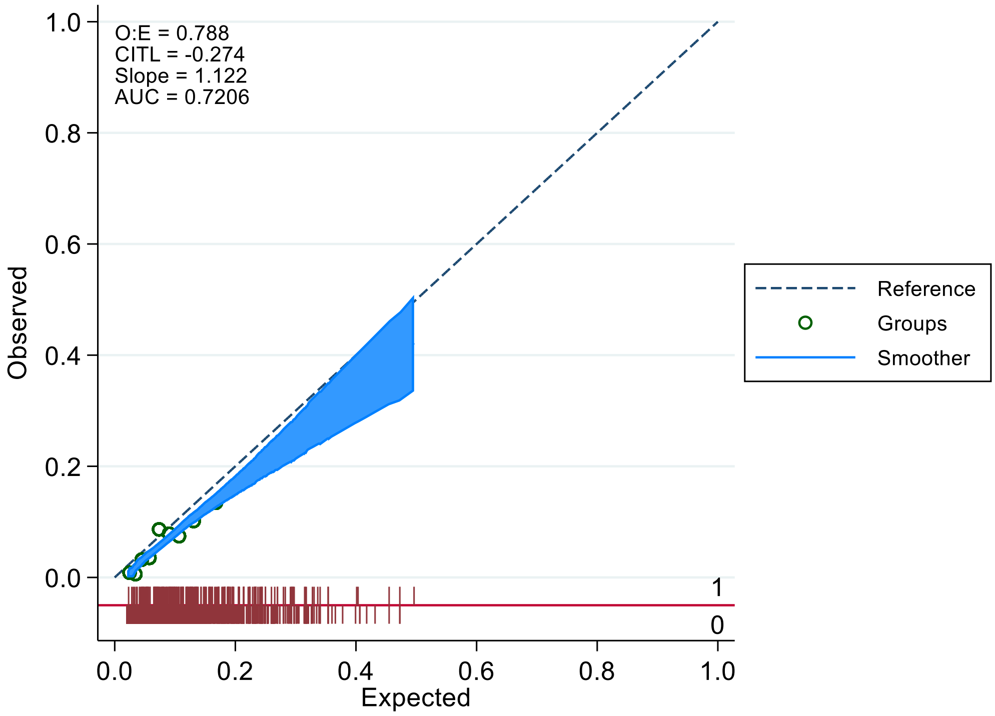

**Legend:** Reference (dashed line) indicates perfect calibration; Lowess (blue line) indicates calibration slope; Red rug plots indicate the distribution of predicted risk of participants with hypoxemia (above the line), and those without hypoxemia (below the line). E:O = expected to observed events ratio; CITL = calibration-in-the-large; AUC = area under the receiver operating characteristic curve.
